# Timing of selective serotonin reuptake inhibitor use and risk for preterm birth and related adverse events

**DOI:** 10.1101/2023.03.03.23286717

**Authors:** Yeon Mi Hwang, Ryan T. Roper, Samantha N. Piekos, Daniel A. Enquobahrie, Mary F. Hebert, Alison G. Paquette, Priyanka Baloni, Nathan D. Price, Leroy Hood, Jennifer J. Hadlock

## Abstract

**Purpose:** There is uncertainty around the safety of SSRIs for treating depression during pregnancy. We aimed 1) to address confounding by indication, as well as socioeconomic and environmental factors associated with depression and 2) evaluate associations of timing of SSRI exposure in pregnancy with the risk of preterm birth and related outcomes (small for gestational age and low birthweight) among women with depression before pregnancy.

**Methods:** We conducted propensity score-adjusted regression to calculate odds ratios (OR) of preterm birth, small for gestational age, and low birth weight. We accounted for maternal/pregnancy characteristics, pre-pregnancy comorbidity/depression severity, social vulnerability, rural health disparity, and pre-natal depression severity. We additionally conducted a drug-specific analysis and assessed the impact of other classes of antidepressants within our cohort of interest.

**Results:** Among women with a history of depression, we identified women with indication of depression ≤ 180 days before pregnancy (n=6,408). Women with no SSRI order during pregnancy (n=3,122) constituted the unexposed group (no SSRI exposure group). The late SSRI exposure group consisted of women with an SSRI order after the first trimester (n=2,596). The early-only SSRI exposure group consisted of women with SSRI orders only in the first trimester (n=691). Late SSRI exposure group had an increased risk of preterm birth of OR=1.7 ([1.3,2.2], p<0.0001), and low birth weight of OR = 1.7 ([1.3,2.4], p<0.001), relative to the no SSRI exposure group.

**Conclusions:** These findings suggest associations between preterm birth/low birthweight and SSRI exposure is dependent on exposure timing during pregnancy. Small for gestational age is not associated with SSRI exposure.

## Introduction

Depression is one of the most common comorbidities women experience during pregnancy or the postpartum period, occurring at a rate of 10-20% in the US from 2004 to 2012^1^.

Besides, Severe Acute Respiratory Syndrome Coronavirus (COVID-19) Pandemic has significantly increased the risk of maternal depression^2^ and common symptoms of long COVID include depression^3^. Between 2004 and 2008, 8% of pregnant women took antidepressants^4–7^. As the rate of maternal depression is increasing and some studies ^8,9^ suggest potentially reduced risk of COVID-19 severity associated with SSRIs, we suspect more prevalent use of antidepressants among pregnant patients. Despite the high prevalence of prenatal antidepressant use, its safety during pregnancy is not fully understood. There is little agreement on the association between antidepressants and preterm birth (PTB; birth prior to 37 weeks gestation). PTB is a key contributor to perinatal morbidity and mortality in developed and developing countries and occurs in 10% of all live births in the US ^10,11^.

Although studies of varying size and design have reported results supporting the association between antidepressant use and PTB risk^6,12–16^, findings have been inconsistent, and inadequate control for underlying depression has raised concerns of potential confounding-by-indication^17,18^. Additionally, PTB risk may be influenced by the timing of antidepressant exposure^6,12,13,19,20^. Further, uncertainty around the safety of prenatal antidepressant use led 75% of women to discontinue antidepressants before the second trimester of pregnancy, according to a cohort study of 228,876 singleton, Medicaid-covered pregnancies in Tennessee between 1995 and 2007^6^. SSRIs are the most common class of antidepressants^6,13^. The objective of this study is to evaluate the risk of PTB and other relevant adverse outcomes, low birthweight (LBW) and small for gestational age (SGA), following SSRI use during the second or third trimester. To minimize confounding-by-indication, we conduct the study among women with depression onset before the pregnancy. We comprehensively addressed the influences of confounding factors, including not only pregnancy characteristics and comorbidities but also environmental factors based on the geographical region the patient resides. We also characterize the influence of timing of SSRI intake by distinguishing the SSRI exposure groups based on exposure status after the first trimester. We also conducted SSRI drug-specific analyses and additional analyses on the other classes of antidepressants. We

## Methods

### Study Setting and Study Population

We conducted a retrospective study among women who delivered between 2010/01/01 and 2020/12/31 (n=436,796) at Providence St. Joseph Health (PSJH) across Alaska, California, Montana, Oregon, and Washington.

Figure 1 shows the study population selection. We selected mothers who were continuously enrolled from 180 days before the last menstrual period (LMP) to the date of delivery. We confirmed the date of delivery was at least 20 weeks apart from the LMP and ‘living’ delivery status of the baby. Since multiple gestation is a significant risk factor for PTB, we limited our cohort to singleton pregnancies. We excluded deliveries prior to 2013 because the EHR was not used broadly for maternity across PSJH. From this source population (n=284,436), women with depression onset before pregnancy were selected to avoid confounding-by-indication (n=6,743). We first identified women diagnosed with depression any time before the pregnancy. From this population of women with a history of depression, we selected women with a diagnosis of depression or any antidepressant prescription order during the six-month pre-pregnancy period (LMP-180 days∼LMP). Women with bupropion or trazodone prescriptions and no other antidepressants were excluded unless they had a diagnosis of depression because these medications are often prescribed as smoking cessation medications or sleep aids^21,22^. We excluded women with a diagnosis of psychosis or bipolar disorder during the two-year pre-pregnancy period (LMP-730 days∼LMP) and fetal sex of ‘other’ category. The final analytic population consisted of 6,408 women.

**Figure 1.**
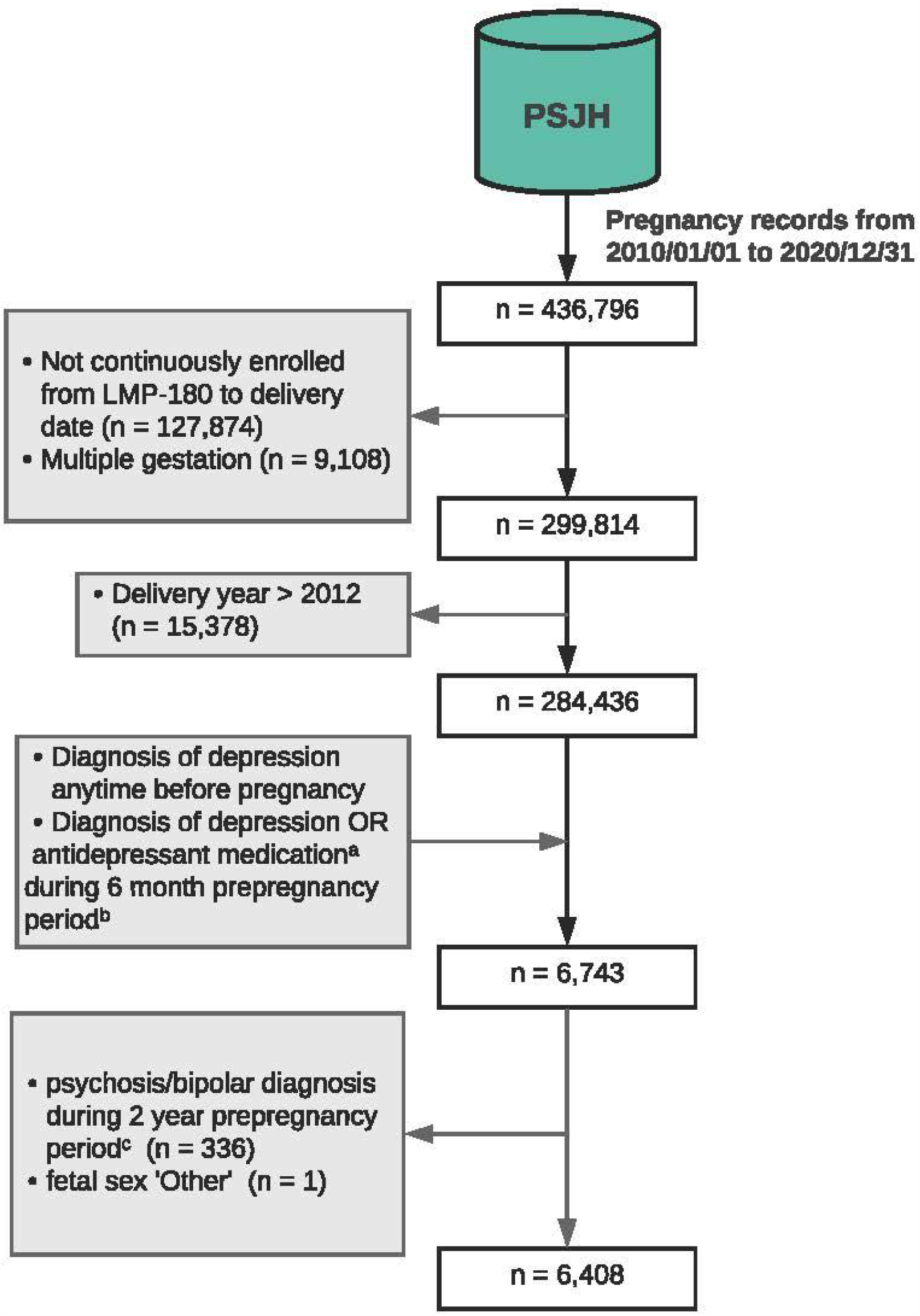
Flow diagram of cohort selection. Abbreviations: EHR, Electronic Health Record; LMP, Last Menstrual Period - the start of the pregnancy; PSJH, Providence St. Joseph Hospital. a. women with only bupropion or trazodone order were excluded b.6-month pre-pregnancy period is the time frame between 180 days before the LMP to LMP c. two-year pre-pregnancy period is the time frame between 730 days before the LMP to LMP Inclusion criteria are indicated with arrows toward the flow arrow. Exclusion criteria are indicated with arrows away from the flow arrow.

### Measures

#### Outcome

Our primary outcome was PTB, determined by gestational age at birth (GA; GA<37 weeks). Gestational age at birth (GA) was a proxy outcome of PTB, calculated based on the expected date of delivery and actual delivery date. Secondary outcomes were LBW (birthweight < 2,500g) and SGA (birthweight < 10th percentile of the same gestational age).

#### Exposure

The main exposure of interest was SSRI exposure after the first trimester (GA≥ 13 weeks). The secondary exposure of interest was SSRI exposure only in the first trimester (Figure 2, Figure S1). Study participants were divided into three groups based on SSRI exposure status during pregnancy. The control group (no SSRI exposure) was women with no SSRI prescribed during the pregnancy. The late SSRI exposure group was women who had any SSRI order after the first trimester. The early-only exposure group was of women with SSRI orders only in the first trimester.

**Figure 2.**
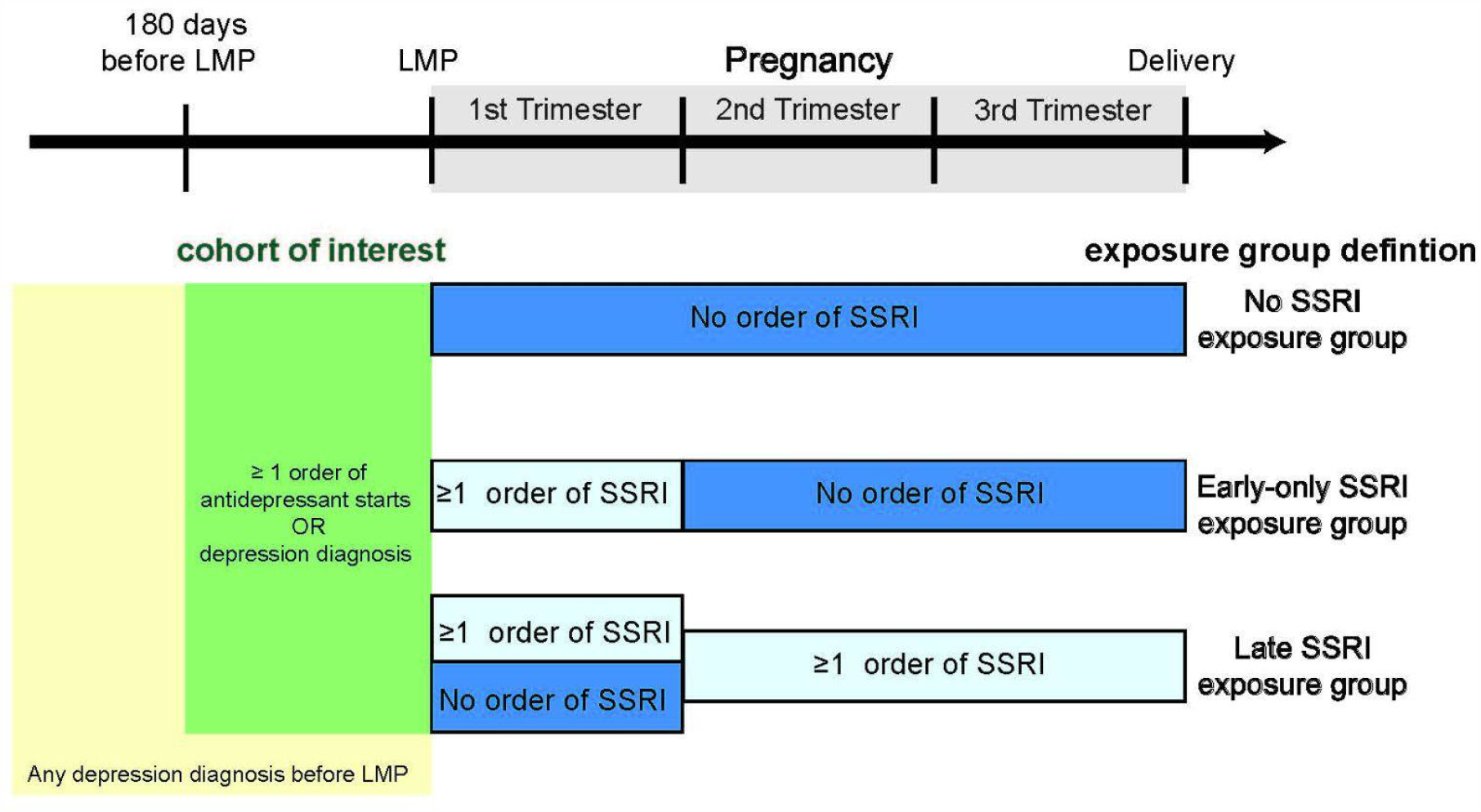
Study Design. Abbreviations: LMP, Last Menstrual Period; SSRI, Selective Serotonin Reuptake Inhibitor Depressed cohort is subdivided into no, early-only, and late SSRI exposure groups. No SSRI exposure group has no order of SSRI during pregnancy. Early-only SSRI exposure group has SSRI order only at the first trimester, but none at the second or third trimester. Late SSRI exposure group has any order during the second or third trimester.

#### Covariates

We extracted information on maternal characteristics, pre-pregnancy/prenatal mental conditions, and pre-pregnancy comorbidities from the EHR (Table S1). Pregnancy and maternal characteristics were collected during prenatal care or at the time of delivery. These included parity, preterm history, delivery year, fetal sex, age at LMP, self-reported racial group, self-reported ethnic group, insurance, pregravid body mass index (BMI) category, self-reported smoking status, self-reported illegal drug use status, self-reported ethanol consumption status, and Centers for Disease Control and Prevention social vulnerability index (CDC-SVI). CDC-SVI represents percentile ranking of each census tract on 15 social factors. Social factors themes include socioeconomic status, household composition, race/ethnicity/language, and housing/transportation^23^. Pre-pregnancy mental conditions were determined based on prescription orders (Table S1), clinical diagnoses (Table S1), and Patient Health Questionnaire-9 (PHQ-9) scores ^24,25^. We collected pre-pregnancy exposure status of N06AA (non-selective monoamine reuptake inhibitor), N06AB (SSRI), N06AF (monoamine oxidase inhibitors, non-selective), N06AG (monoamine oxidase A inhibitors), and N06AX (other antidepressants) during the six months prior to the pregnancy. We extracted diagnoses of adjustment disorders and anxiety disorders from the two-year pre-pregnancy period. Prenatal mental health was determined based on depression diagnosis and PHQ-4 score during late pregnancy^26,27^. Other comorbid diagnoses (anemia, asthma, cardiovascular diseases, cystic fibrosis, chronic lung diseases, diabetes, renal diseases, leukemia, pneumonia, sepsis, and sickle cell diseases) were included when active during the two-year pre-pregnancy period (Table S1).

### Statistical analyses

#### Descriptive statistics

Descriptive statistics for maternal characteristics, outcomes, covariates, prenatal mental conditions are presented by exposure group (Table 1). We examined the distribution of covariates across exposure groups (no, early-only, late SSRI exposure group) and outcome groups (PTB and non-PTB). We used contingency chi-squared test for categorical variables. For continuous variables, we used one-way ANOVA test and Welch’s t-test. Additionally, we performed multiple comparison post hoc correction using the Benjamin-Hochberg procedure for comparisons on combinations of exposure groups.

**Table 1.**
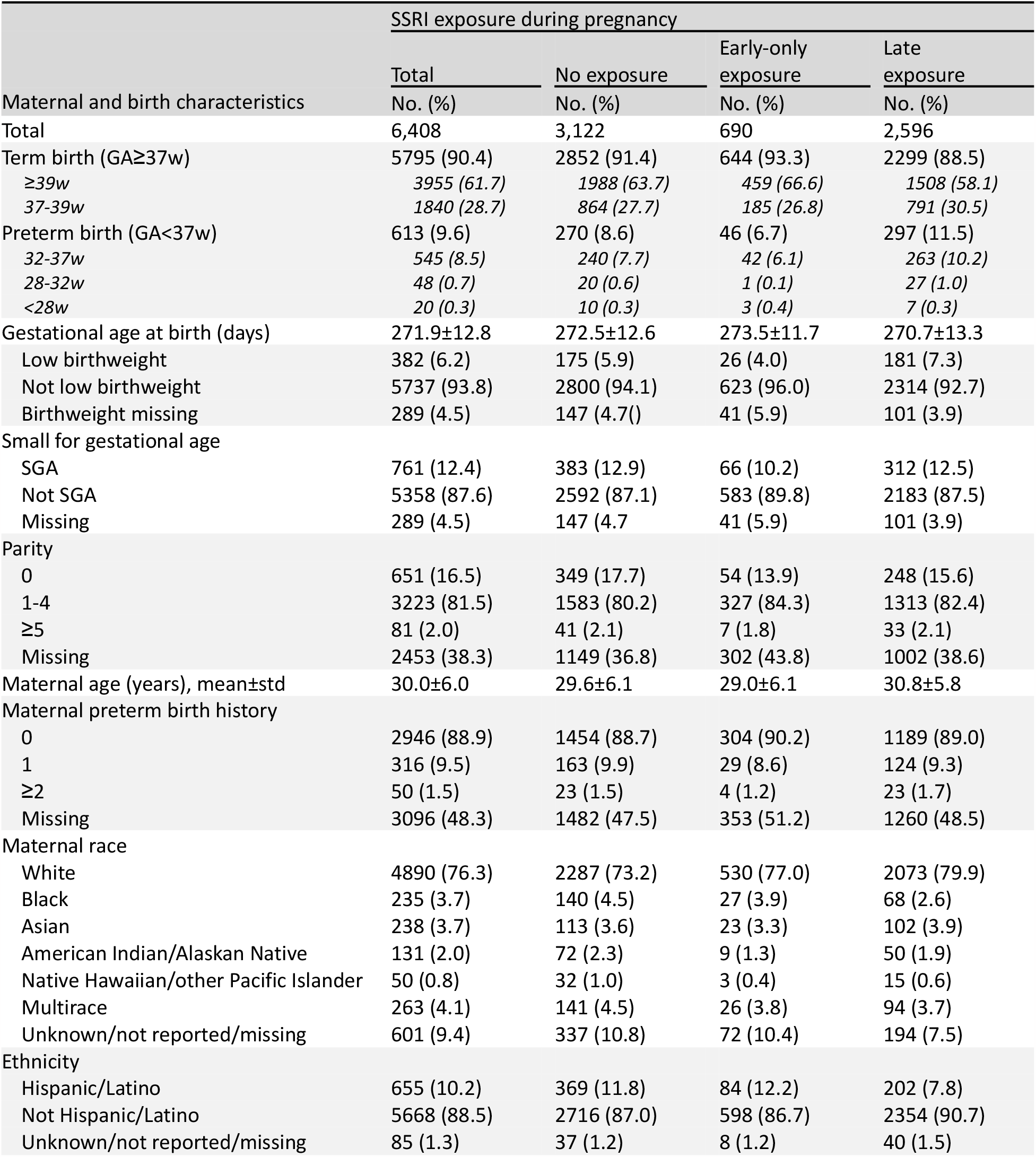
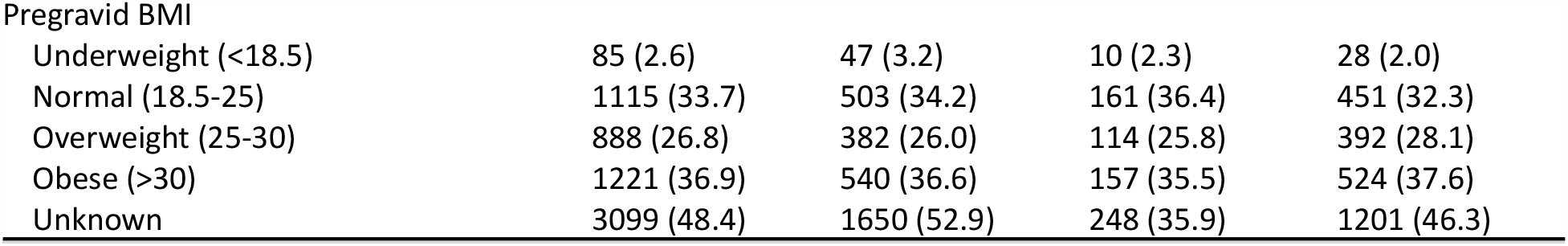
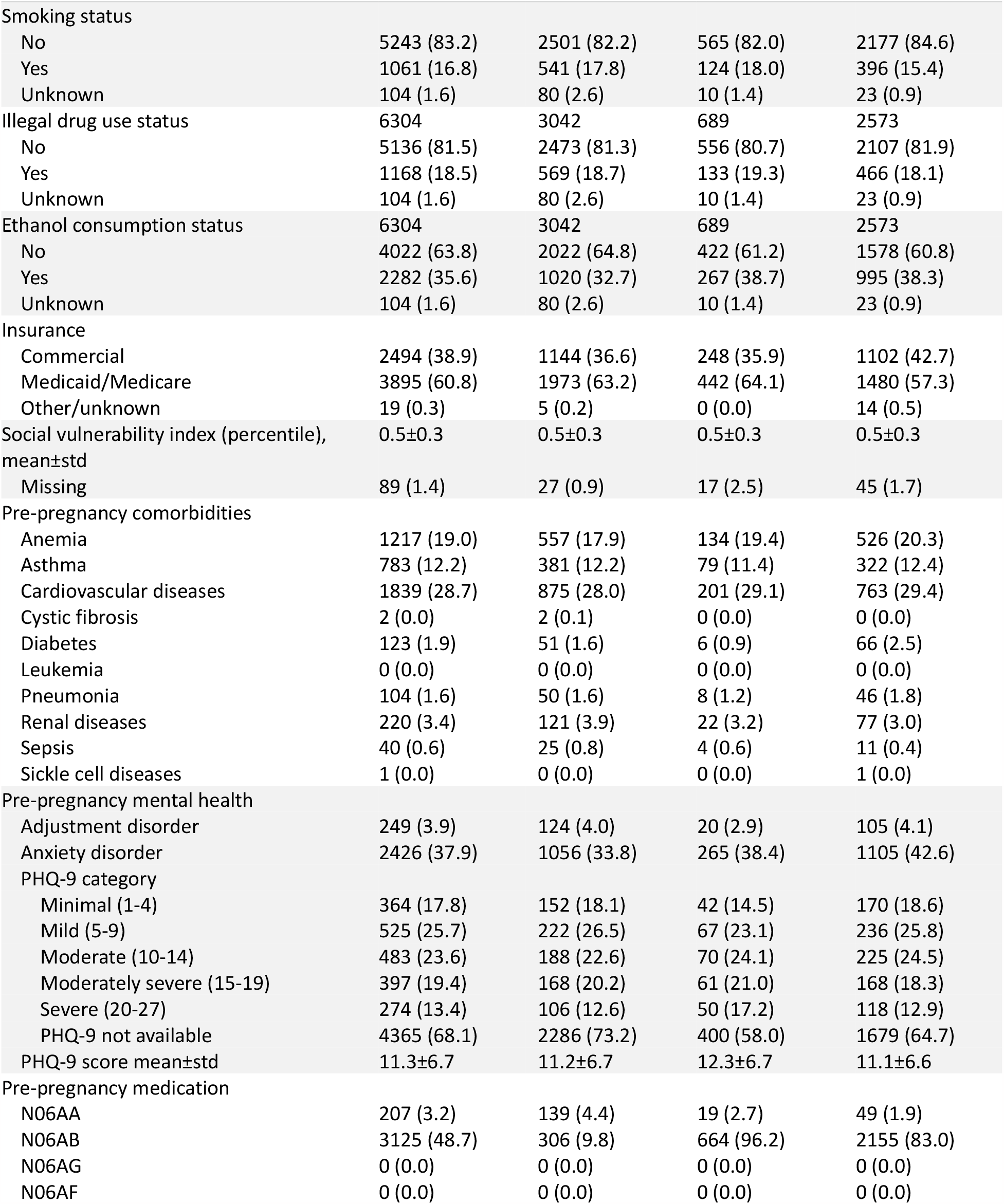
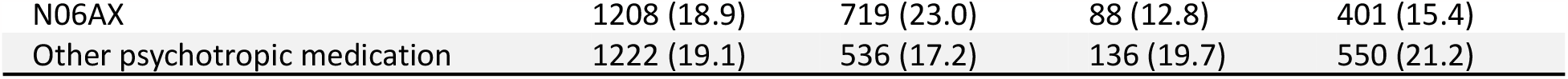
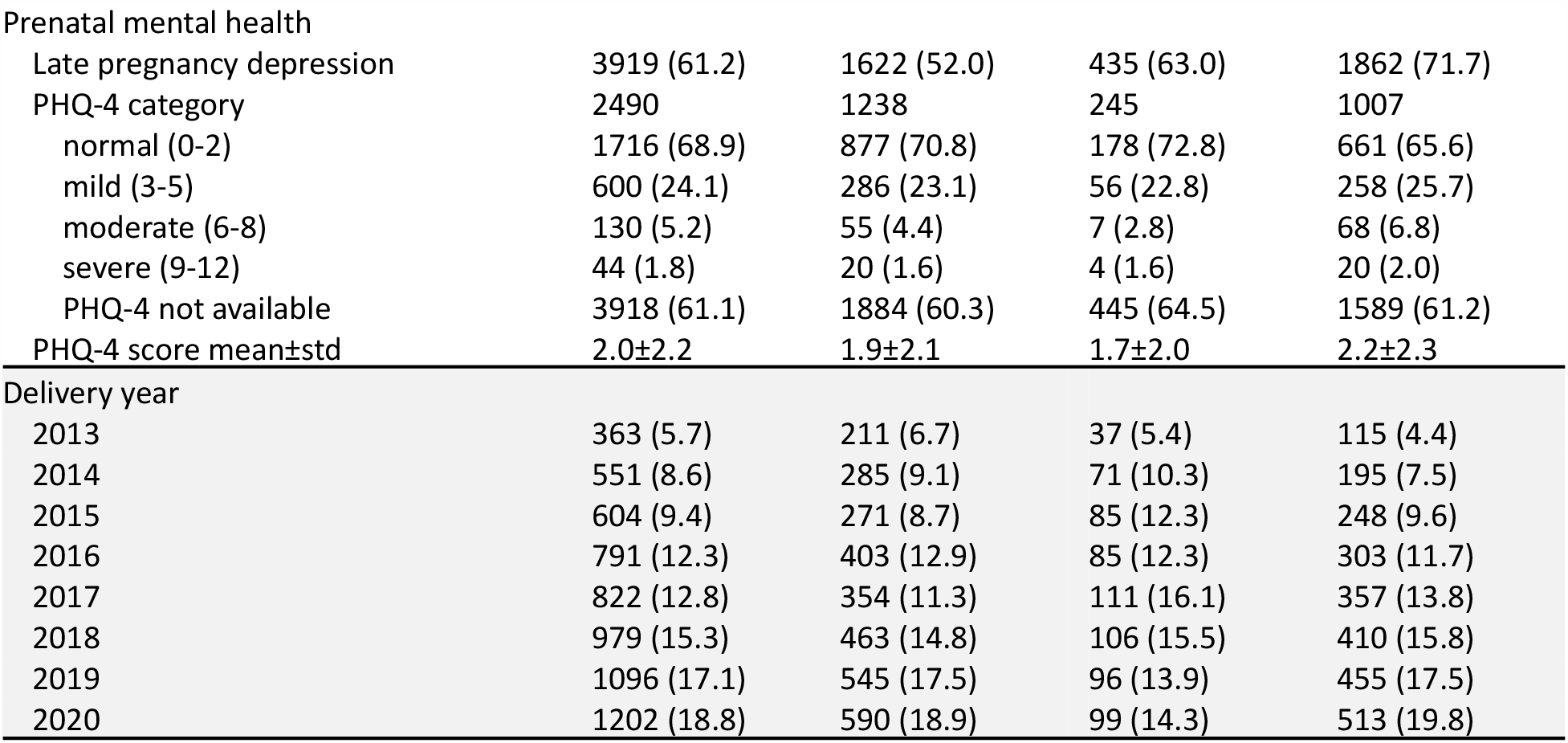
Descriptive statistics for women who had depression onset before pregnancy who went on to deliver, by SSRI exposure. For categorical variables, the number of samples and proportion (%) in each exposure group are reported. For continuous variables, mean and standard deviation were reported. Abbreviations: BMI, Body Mass Index; LMP, Last Menstrual Period; PHQ, Patient Health Questionnaire; SSRI, Selective Serotonin Reuptake Inhibitor, N06AA (non-selective monoamine reuptake inhibitor), N06AB, SSRI); N06AF, monoamine oxidase inhibitors, non-selective; N06AG, monoamine oxidase A inhibitors; N06AX, other antidepressants; std, standard deviation. All categorical data are presented as n (%). All continuous data are presented as mean±standard deviation. All variables are defined in Table S

#### Main analysis

We used propensity score adjusted log-binomial and linear regression to calculate the odds ratio (OR) of PTB, LBW, and SGA, and the difference in the mean of GA corresponding to exposure to SSRI. We applied multivariable imputation for missing values for parity, PTB history, race, engraved BMI, smoking status, illegal drug use status, ethanol consumption status, and CDC-SVI ^28^. For each comparison, we selected the treatment group and calculated the propensity score using logistic regression. Covariates of the main model included pregnancy and maternal characteristics, pre-pregnancy mental conditions, and pre-pregnancy comorbid conditions as defined above.

We compared the late SSRI exposure group to the non-late, early-only, and no SSRI exposure group (Figure S1). We compared the early-only SSRI group to the no SSRI exposure group. Additionally, any SSRI exposure group was compared with the no SSRI exposure group (Figure S1). These associations were further assessed as follows. First, we subdivided the late SSRI exposure group into late-only and both SSRI exposure groups to evaluate the effect of late-only SSRI exposure (Figure S1). Both SSRI exposure groups consisted of women who had SSRI orders during and after the first trimester.

Late-only SSRI exposure group consisted of women who had SSRI order only after the first trimester (Figure S1). Second, drug-specific analyses were conducted on individual SSRI medications: citalopram, escitalopram, fluoxetine, paroxetine, sertraline and fluvoxamine.

#### Sensitivity Analysis

First, the influence of late SSRI exposure was evaluated among women who had depression diagnoses at the late pregnancy (n=3,928). This approach was adopted to adjust for the prenatal depression severity. We assumed women with severe prenatal depression symptoms were likely to have depression diagnosis during the pregnancy. Second, to assess the impact of pre-pregnancy depression severity, we did a sensitivity analysis on a subsample of women who took PHQ-9 during the two-year pre-pregnancy period (n=2,043). In this analysis, we included the most recent PHQ-9 score as a covariate within the main model. Similarly, we assessed the impact of the prenatal depression severity using the Patient Health Questionnaire-4 (PHQ-4) score^26,27^. Fourth, we conducted an analysis by excluding women who were exposed to other categories of antidepressants (N06AA, N06AG, N06AF, N06AX). Fifth, we added additional psychiatric medication as a covariate to the main model to examine the potential impact of other psychiatric medications to the risk of PTB, SGA, and LBW. Finally, we ran a sensitivity analysis restricted to women with more than one prescription during pregnancy as women prescribed with SSRI might not necessarily take it.

#### Additional Analysis

In addition, we examined the effect of other antidepressants (N06AA, N06AG, N06AF, N06AX) exposure on PTB, SGA, LBW. This approach was to further distinguish associations resulting from SSRI and other antidepressant exposures. We additionally separated Serotonin and norepinephrine reuptake inhibitor (SNRI) from N06AX, as it is the most common class of N06AX antidepressants.

## Results

### Descriptive statistics

Table 1 presents the descriptive statistics of covariates, outcomes, and maternal mental health. Of 284,436 women, 6,408 women were eligible as our cohort of interest (Study Participants, Figure 1). The study cohort was divided into three groups (Figure 2, Figure S1): 3,122 women in no SSRI exposure group, 690 women in early-only SSRI exposure, and 2,596 women in late SSRI exposure. Among these three exposure groups, the late SSRI exposure group had the highest rate of PTB (11.5%), the no SSRI exposure had a rate of 8.6%, and the early-only SSRI exposure group had the lowest PTB prevalence rate (early-only: 6.7%) Likewise, the late SSRI exposure group had the shortest, and the early-only SSRI exposure group had the longest GA (mean GA±standard deviation; no: 272.5±12.6, early-only: 273.5±11.7, late: 270.7±13.3).

Among the subpopulation with pre-pregnancy PHQ-9 score (n=2,043), pre-pregnancy PHQ-9 score was the lowest for the late SSRI exposure group and highest for the early-only SSRI exposure group (no: 11.2±6.7, early-only: 12.3±6.7, Late: 11.1±6.6; P < 0.05) (Table 1, Table S2). In contrast, among the subpopulation with prenatal PHQ-4 score (n=2,490), the PHQ-4 score was the highest for the late SSRI exposure group and lowest for the early-only SSRI exposure group (no: 1.9±2.1, early-only: 1.7±2.0, late: 2.2±2.3, P < 0.05) (Table 1, Table S2). Although pre-pregnancy PHQ-9 score and prenatal PHQ-4 score were significantly different across exposure groups, they were not significantly different across outcome groups (P=0.8, P=0.2; Table S2)

### Main analysis

#### Increased risk of preterm birth and low birthweight for late SSRI exposure

Late SSRI exposure was associated with elevated risk of PTB, LBW, and shortened GA after controlling for confounders (Figure 3, Table S3). The odds ratios (OR) of PTB were 1.6 ([1.3,1.9], P<0.0001), 1.7 ([1.3,2.2], P<0.0001) and 1.5 ([1.1,2.2], P<0.05) and those of LBW were 1.7 ([1.3,2.2], P<0.0005), 1.7 ([1.3,2.4], P<0.0005), and 1.6 ([1.1,2.5], P<0.05) when comparing late SSRI exposure group with no-late, no, and early-only SSRI exposure groups, respectively. GA was significantly decreased by 2.3 days ([-3.1,-1.5], P<0.0001), 2.6 days ([-3.6,-1.5], P<0.0001), and 1.9 days ([-3.1,-0.8], P<0.005) in the late SSRI exposure group compared to the no-late, no, and early-only SSRI exposure groups, respectively.

**Figure 3.**
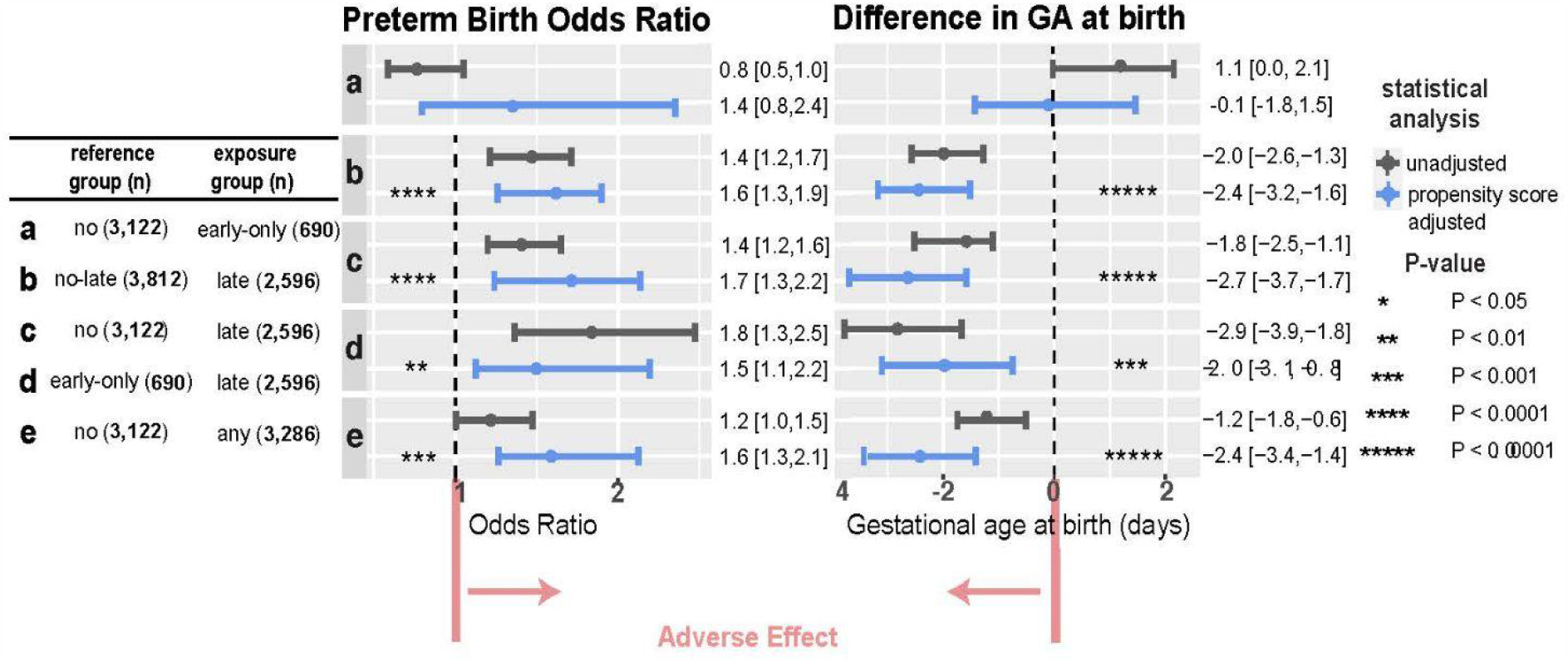
Increased risk of preterm birth and decreased gestational days at birth for women exposed to SSRI in second or third trimester. Abbreviations: CI, Confidence Interval; GA, Gestational Age. The odds ratio of preterm birth and difference in the mean of GA at birth were calculated using log-binomial regression and linear regression adjusting for propensity score. Exposure groups were defined as follows. No: women with no SSRI exposure during pregnancy. Early only: women with SSRI exposure only in the first trimester. Late: women with SSRI exposure in the second or third trimester. Any: women with any SSRI exposure during pregnancy (early only + late). No-late: women with no SSRI exposure in the 2nd nor 3rd trimester (no + early only). Each comparison (a-e) is defined in the table on the left. +: P < 0.1; *: P < 0.05; **: P < 0.01; ***: P < 0.001; ****: P < 0.0001; *****: P < 0.00001

Early-only SSRI exposure was not associated with either PTB (OR=1.4, [0.8,2.4]), LBW (OR=1.0, [0.5,2.0]), or shortened length of gestation (β =-0.1, [-1.8,1.5]). Women exposed to SSRI at any time during pregnancy had 1.6 ([1.2,2.0], P<0.0005) fold higher risk of PTB, 1.6 ([1.2,2.2], P<0.001) fold higher risk of LBW, and delivered 2.3 ([-3.2,-1.3], P<0.0001) days earlier than the unexposed to SSRI group (Figure 3, Table S3). Most (both SSRI exposure group, n=2,259, 87.0%) of the late SSRI exposure group were taking SSRI throughout the pregnancy, based on reports in EHR data. The late-only SSRI exposure group (n=337) had similar PTB and LBW risk (PTB OR=1.1 [0.7,1.8]; LBW OR=0.9 [0.5,1.7]) and mean GA (β=-0.3 [-2.5,1.8]) compared with both SSRI exposure group (Table S4).

### Drug-specific Analysis

In relation to PTB, drug-specific analyses on fluoxetine and sertraline demonstrated consistent results as the main SSRI analysis and remained statistically significant. Whereas, citalopram, escitalopram, and paroxetine showed inconsistent results (Table S5). There was increased risk of PTB with early-only exposure to citalopram (OR=2.1 [1.0,4.1], P<0.05) and escitalopram (OR=2.4 [1.0,5.7], P<0.05) when compared to their no exposure groups (Table S5). In relation to LBW, all drugs (citalopram, escitalopram, fluoxetine, paroxetine, and sertraline) showed similar result as the main SSRI analysis. No woman was exposed to fluvoxamine during pregnancy; thus fluvoxamine was excluded from the drug-specific analysis.

### Sensitivity analysis

#### Adjustment for pre-pregnancy and prenatal depression severity

The strong and significant association between late SSRI exposure and PTB/LBW persisted when pre-pregnancy and prenatal depression severities were addressed (Table S6). In the pre-pregnancy depression severity addressed model, the late SSRI exposure group still had a higher chance of PTB (OR=1.4 [1.0,1.9], P=0.05) and LBW (OR=1.8 [1.0,3.2], P<0.05) compared to women with equivalent severity of depression but no SSRI exposure (Table S6). When prenatal depression severity was adjusted, the late SSRI exposure group had increased risk of LBW (OR=1.6 [1.1, 2.5], P<0.01), borderline statistically significant increased risk for PTB (OR=1.3 [0.9,1.9], P=0.11) relative to no SSRI exposure (Table S6).

## Discussion

In this study, we examined the relationship between the timing of prenatal SSRI exposure and three outcomes (PTB, LBW and SGA) among women with a history of depression and adjusted for confounders that have been linked to PTB, including parity, preterm history, maternal mental health, and comorbidities. We found that women prescribed SSRIs in late pregnancy were 1.7 times more likely to deliver preterm when compared to women who were not prescribed SSRIs. Our findings were robust to several sensitivity analyses that accounted for depression severity and exposure to other categories of antidepressants.

Although SSRI exposure in late pregnancy was associated with increased PTB risk and discontinuation in early pregnancy was not, drug-specific analysis revealed that the association was dependent on the type of SSRI. Sertraline, the most commonly prescribed SSRI, and fluoxetine followed the pattern of SSRI. Citalopram and escitalopram were the opposite. Discontinuation in early pregnancy was associated with an increased risk of PTB, and exposure in late pregnancy was not.

Our findings are consistent with several prior observational studies investigating the timing of prenatal antidepressant or SSRI-specific exposure and PTB^6,12,14–16^. A systematic review and meta-analysis of 41 observational studies reported that second and third trimester antidepressant exposure increased the risk of PTB^12^; 22 (53.6%) of studies examined SSRI exposure. A retrospective study on a similar-sized singleton pregnancy cohort covered by Tennessee Medicaid also suggested the association between second trimester antidepressant (SSRI or non-SSRI antidepressant) exposure and PTB, but not with first trimester exposure^6^.

In contrast with our study, some studies reported an elevated risk for PTB for first trimester SSRI or antidepressant (non-SSRI antidepressant) exposure^13,19,20^. However, the first trimester exposure in these studies included women who continued the use throughout the pregnancy. Thus, the gestational timing of SSRI exposure on PTB risk cannot be determined in these studies. A population-based cohort study conducted in Finland reported lower risk of PTB for women who purchased SSRIs during pregnancy; the risk of PTB was 16% lower even when accounting for the underlying depression^29^. However, that study compared women exposed to SSRI to unexposed ones with psychiatric diagnosis during the pregnancy. Our study compared only those participants experiencing depression before the pregnancy.

Surprisingly, pre-pregnancy depression severity did not appear to be relevant to the risk of PTB, based on PHQ-9 score. These outcomes are contrary to earlier studies suggesting preconception stress and mental health correlation to PTB^17,30,31^. However, we restricted our study to include participants with depression onset prior to the pregnancy, while those studies do not have such restrictions. Presence of pre-pregnancy depression may be a more significant contributor to PTB than severity of depressive symptoms. One possible explanation for this result is the saturation of maternal stress-induced response. Maternal stress may induce a response, such as hypothalamic-pituitary adrenal (HPA) axis dysfunction^32,33^ or inflammatory immune response^34^ which activates a mechanism leading to PTB and this response saturates and plateau once stress hits the level of depression.

### Strengths and Limitations

In this study, we minimized confounding-by-indication by analyzing women with depression onset prior to pregnancy. All study participants had a history of depression and had indication of depressive symptoms (antidepressant prescription or depression diagnosis) within the 6-month pre-pregnancy period. We stratified the exposure group based on the late pregnancy exposure status to represent the mother’s SSRI usage patterns. In our study, among women with depression onset before the pregnancy, 40.4% of women chose to take SSRI in the late pregnancy and 10.8% discontinued during the first trimester.

We analyzed a large sample to provide sufficient statistical power to examine PTB, SGA, and LBW. We had 299,814 singleton pregnancy delivery records in the PSJH database and 6,408 samples in our analytic cohort. 613 (9.6%) patients in this analytic cohort delivered preterm. This allowed observation of statistically significant results in subgroup analyses and sensitivity analyses.

We applied rigorous sensitivity analyses to test the robustness of associations observed in the main analysis. We accounted for depression severities before and during the pregnancy and exposure to antidepressant polytherapy. Many prior studies on the association between SSRI and PTB were limited due to lack of severity data ^6,16,19,35–39^.

We subdivided late exposure group into late-only and both (early and late) exposed group in order to address the potential concern that increased PTB risk of late exposed group was driven by the cumulative SSRI exposure, rather than some condition specific to late pregnancy. However, the late-only SSRI exposure group had a similar risk of PTB when compared to the both-exposed group, indicating late SSRI exposure drives the association with increased risk of PTB.

We included the CDC social vulnerability index as a covariate. CDC social vulnerability index accounts for 15 social factors on socioeconomic status, household compositions, race/ethnicity/language, and housing/transportation. CDC uses U.S. Census data to calculate social vulnerability of each census tract, a subdivision of counties ^23^. This index allows us to account for social disparity based on the location of residence. To our knowledge, this is the first study that accounted for the social vulnerability when assessing the impact of SSRI to risks of adverse pregnancy outcomes.

Additionally, we assessed SSRI drug-specific associations and N06AA/N06AX associations with PTB, SGA, and LBW. This helps us understand the magnitude of individual drug association with PTB,SGA, and LBW and to discern the contributions of other antidepressants (N06AA/N06AX). We also assessed class-specific association between SNRI and outcomes as SNRI is the major class of N06AX ATC category.

A potential weakness of this study was the potential misclassification of exposure. Prescription orders do not necessarily indicate actual exposure. Non-differential misclassification of exposure biases toward the null, which could potentially attenuate the strength of the association. This is an inherent limitation in EHR data, but the risk misclassification is somewhat mitigated because the study population is pregnant women; prenatal visits have short cycles and medication reconciliation is an integral part of prenatal visits ^40^.

Another potential limitation of this study was immortal time bias induced from the exposure time period of the late SSRI exposed group. The later onset of the exposure leads to longer immortal time, in which the outcome cannot occur ^41^. We attempted to minimize the immortal time bias by selecting women who delivered living birth in ≥ 20 GA weeks as the source population.

We did not distinguish between spontaneous PTB and medically indicated PTB. We had insufficient information and resources to accurately categorize PTB based on the subphenotype, as that information was recorded in the format of unstructured free-text notes. We plan to replicate this study focusing on spontaneous PTB once we have the resource to apply Natural Language Processing (NLP) technique to transform unstructured data into structured data.

We focused the outcome of interest specifically on PTB and related outcomes, LBW and SGA. SSRI may have a protective impact on adverse maternal outcomes. Future studies will investigate the impact of SSRI on other adverse maternal outcomes, such as miscarriage and stillbirth. Additionally, we considered each delivery record as an independent entity, ignoring the correlations of data points coming from the same individual. In the future, if sample size allows, we may replicate this study in a subpopulation of primaparae to validate our results.

Depression severity assessments, PHQ-9 and PHQ-4 are not administered to all pregnant women. Although we were able to perform sensitivity analyses on subpopulation of people who took the assessments, we had insufficient number of samples to perform stratified analyses based on the depression severity. SSRI may have differential effects on women with more severe depressive symptoms ^42,43^. Going forward, stratified analysis based on pre-pregnancy and prenatal depression severity should be conducted to further evaluate the impact of SSRI on PTB.

### Conclusion

Among women with depression onset prior to pregnancy, SSRI exposure after the first trimester is associated with an increased risk for preterm birth and low birthweight.

Although our findings suggest that there is an association between SSRI exposure after the first trimester of pregnancy and risk of PTB and LBW, depression should not be left untreated during the pregnancy. Untreated depression increases the chance of adverse pregnancy outcomes and high-risk health behaviors ^44,45^. Both patients and physicians should be informed on both the risk and benefit of medication treatment during pregnancy to make optimal patient specific decisions given the patient’s health conditions.

## Data Availability

Results have been aggregated and reported within this paper to the extent possible while maintaining privacy from personal health information as required by law. Data are archived within Providence St Joseph Health systems in a HIPAA-secure audited compute environment.

## Acknowledgments

We thank PSJH for sharing their data engineering expertise and computational resources. We acknowledge SNOMED International for developing and maintaining SNOMED-CT©.

## Figure legends

**Figure S1:** Flow diagram of exposure group selection Abbreviation: SSRI, Selective Serotonin Reuptake Inhibitor.

## References

[1] Van Niel MS, Payne JL. Perinatal depression: A review. Cleve Clin J Med. 2020;87(5):273–277.

[2] Hessami K, Romanelli C, Chiurazzi M, Cozzolino M. COVID-19 pandemic and maternal mental health: a systematic review and meta-analysis. J Matern Fetal Neonatal Med. Published online November 1, 2020:1–8.

[3] Kyzar EJ, Purpura LJ, Shah J, Cantos A, Nordvig AS, Yin MT. Anxiety, depression, insomnia, and trauma-related symptoms following COVID-19 infection at long-term follow-up. Brain Behav Immun Health. 2021;16:100315.

[4] Mitchell AA, Gilboa SM, Werler MM, Kelley KE, Louik C, Hernández-Díaz S, et al. Medication use during pregnancy, with particular focus on prescription drugs: 1976-2008. Am J Obstet Gynecol. 2011;205(1):51.e1-e8.

[5] Huybrechts KF, Palmsten K, Mogun H, Kowal M, Avorn J, Setoguchi-Iwata S, et al. National trends in antidepressant medication treatment among publicly insured pregnant women. Gen Hosp Psychiatry. 2013;35(3):265–271.

[6] Hayes RM, Wu P, Shelton RC, Cooper WO, Dupont WD, Mitchel E, et al. Maternal antidepressant use and adverse outcomes: a cohort study of 228,876 pregnancies. Am J Obstet Gynecol. 2012;207(1):49.e1-e9.

[7] Andrade SE, Raebel MA, Brown J, Lane K, Livingston J, Boudreau D, et al. Use of antidepressant medications during pregnancy: a multisite study. Am J Obstet Gynecol. 2008;198(2):194.e1-e5.

[8] Oskotsky T, Maric I, Tang A, Oskotsky B, Wong RJ, Aghaeepour N, et al. Mortality Risk Among Patients With COVID-19 Prescribed Selective Serotonin Reuptake Inhibitor Antidepressants. JAMA Netw Open. 2021;4(11):e2133090.

[9] Nakhaee H, Zangiabadian M, Bayati R, Rahmanian M, Ghaffari Jolfayi A, Rakhshanderou S. The effect of antidepressants on the severity of COVID-19 in hospitalized patients: A systematic review and meta-analysis. PLoS One. 2022;17(10):e0267423.

[10] Rush RW, Keirse MJ, Howat P, Baum JD, Anderson AB, Turnbull AC. Contribution of preterm delivery to perinatal mortality. Br Med J. 1976;2(6042):965–968.

[11] Goldenberg RL, Culhane JF, Iams JD, Romero R. Epidemiology and causes of preterm birth. Lancet. 2008;371(9606):75–84.

[12] Huybrechts KF, Sanghani RS, Avorn J, Urato AC. Preterm birth and antidepressant medication use during pregnancy: a systematic review and meta-analysis. PLoS One. 2014;9(3):e92778.

[13] Sujan AC, Rickert ME, Sara Öberg A, Quinn PD, Hernández-Díaz S, Almqvist C, et al. Associations of Maternal Antidepressant Use During the First Trimester of Pregnancy With Preterm Birth, Small for Gestational Age, Autism Spectrum Disorder, and Attention-Deficit/Hyperactivity Disorder in Offspring. JAMA. 2017;317(15):1553. doi:10.1001/jama.2017.3413

[14] Yonkers KA, Norwitz ER, Smith MV, Lockwood CJ, Gotman N, Luchansky E, et al. Depression and serotonin reuptake inhibitor treatment as risk factors for preterm birth. Epidemiology. 2012;23(5):677–685.

[15] Reis M, Källén B. Delivery outcome after maternal use of antidepressant drugs in pregnancy: an update using Swedish data. Psychol Med. 2010;40(10):1723–1733.

[16] Källén B. Neonate characteristics after maternal use of antidepressants in late pregnancy. Arch Pediatr Adolesc Med. 2004;158(4):312–316.

[17] Phillips GS, Wise LA, Rich-Edwards JW, Stampfer MJ, Rosenberg L. Prepregnancy Depressive Symptoms and Preterm Birth in the Black Women‘s Health Study. Ann Epidemiol. 2010;20(1):8–15.

[18] Li D, Liu L, Odouli R. Presence of depressive symptoms during early pregnancy and the risk of preterm delivery: a prospective cohort study. Hum Reprod. 2009;24(1):146–153.

[19] Lennestål R, Källén B. Delivery outcome in relation to maternal use of some recently introduced antidepressants. J Clin Psychopharmacol. 2007;27(6):607–613.

[20] Ericson A, Källén B, Wiholm B. Delivery outcome after the use of antidepressants in early pregnancy. Eur J Clin Pharmacol. 1999;55(7):503–508.

[21] Wilkes S. The use of bupropion SR in cigarette smoking cessation. Int J Chron Obstruct Pulmon Dis. 2008;3(1):45–53.

[22] Jaffer KY, Chang T, Vanle B, Dang J, Steiner AJ, Loera N, et al. Trazodone for insomnia: A systematic review. Innov Clin Neurosci. 2017;14(7-8):24–34.

[23] CDC/ATSDR SVI Fact Sheet. Published August 30, 2021. https://www.atsdr.cdc.gov/placeandhealth/svi/fact_sheet/fact_sheet.html. Accessed June 28, 2022

[24] Kroenke K, Spitzer RL, Williams JB. The PHQ-9: validity of a brief depression severity measure. J Gen Intern Med. 2001;16(9):606–613.

[25] Levis B, Sun Y, He C, Wu Y, Krishnan A, Bhandari PM, et al. Accuracy of the PHQ-2 Alone and in Combination With the PHQ-9 for Screening to Detect Major Depression: Systematic Review and Meta-analysis. JAMA. 2020;323(22):2290–2300.

[26] Löwe B, Wahl I, Rose M, Spitzer C, Glaesmer H, Wingenfeld K, et al. A 4-item measure of depression and anxiety: validation and standardization of the Patient Health Questionnaire-4 (PHQ-4) in the general population. J Affect Disord. 2010;122(1-2):86–95.

[27] Kroenke K, Spitzer RL, Williams JBW, Löwe B. An Ultra-Brief Screening Scale for Anxiety and Depression: The PHQ–4. Psychosomatics. 2009;50(6):613–621.

[28] Azur MJ, Stuart EA, Frangakis C, Leaf PJ. Multiple imputation by chained equations: what is it and how does it work? Int J Methods Psychiatr Res. 2011;20(1):40–49.

[29] Malm H, Sourander A, Gissler M, Gyllenberg D, Hinkka-Yli-Salomäki S, McKeague IW, et al. Pregnancy Complications Following Prenatal Exposure to SSRIs or Maternal Psychiatric Disorders: Results From Population-Based National Register Data. Am J Psychiatry. 2015;172(12):1224–1232.

[30] Gavin AR, Chae DH, Mustillo S, Kiefe CI. Prepregnancy Depressive Mood and Preterm Birth in Black and White Women: Findings from the CARDIA Study. Journal of Women‘s Health. 2009;18(6):803–811. doi:10.1089/jwh.2008.0984

[31] Grote NK, Bridge JA, Gavin AR, Melville JL, Iyengar S, Katon WJ. A Meta-analysis of Depression During Pregnancy and the Risk of Preterm Birth, Low Birth Weight, and Intrauterine Growth Restriction. Archives of General Psychiatry. 2010;67(10):1012. doi:10.1001/archgenpsychiatry.2010.111

[32] Stephens MAC, Wand G. Stress and the HPA axis: role of glucocorticoids in alcohol dependence. Alcohol Res. 2012;34(4):468–483.

[33] Najafzadeh A. Stress and preterm birth: biological and vascular mechanisms affecting the feto-placental circulation and the length of gestation. Sonography. 2016;3(3):95–102. doi:10.1002/sono.12073

[34] Liu YZ, Wang YX, Jiang CL. Inflammation: The Common Pathway of Stress-Related Diseases. Frontiers in Human Neuroscience. 2017;11. doi:10.3389/fnhum.2017.00316

[35] Oberlander TF, Warburton W, Misri S, Aghajanian J, Hertzman C. Neonatal outcomes after prenatal exposure to selective serotonin reuptake inhibitor antidepressants and maternal depression using population-based linked health data. Arch Gen Psychiatry. 2006;63(8):898–906.

[36] Calderon-Margalit R, Qiu C, Ornoy A, Siscovick DS, Williams MA. Risk of Preterm Delivery and Other Adverse Perinatal Outcomes in Relation to Maternal Use of Psychotropic Medications During Pregnancy. Obstetric Anesthesia Digest. 2011;31(1):46–47. doi:10.1097/01.aoa.0000393180.98348.f6

[37] Lund N, Pedersen LH, Henriksen TB. Selective Serotonin Reuptake Inhibitor Exposure In Utero and Pregnancy Outcomes. Archives of Pediatrics & Adolescent Medicine. 2009;163(10):949. doi:10.1001/archpediatrics.2009.164

[38] Colvin L, Slack-Smith L, Stanley FJ, Bower C. Dispensing patterns and pregnancy outcomes for women dispensed selective serotonin reuptake inhibitors in pregnancy. Birth Defects Research Part A: Clinical and Molecular Teratology. 2011;91(3):142–152. doi:10.1002/bdra.20773

[39] Davis RL, Rubanowice D, McPhillips H, Raebel MA, Andrade SE, Smith D, et al. Risks of congenital malformations and perinatal events among infants exposed to antidepressant medications during pregnancy. Pharmacoepidemiology and Drug Safety. 2007;16(10):1086–1094. doi:10.1002/pds.1462

[40] Kilpatrick SJ, Macones GA, Watterberg KL. Guidelines for Perinatal Care. American Academy of Pediatrics; 2017.

[41] Suissa S. Immortal Time Bias in Pharmacoepidemiology. American Journal of Epidemiology. 2008;167(4):492–499. doi:10.1093/aje/kwm324

[42] Hieronymus F, Lisinski A, Nilsson S, Eriksson E. Influence of baseline severity on the effects of SSRIs in depression: an item-based, patient-level post-hoc analysis. The Lancet Psychiatry. 2019;6(9):745–752. doi:10.1016/s2215-0366(19)30216-0

[43] Fournier JC, DeRubeis RJ, Hollon SD, Dimidjian S, Amsterdam JD, Shelton RC, et al. Antidepressant Drug Effects and Depression Severity. JAMA. 2010;303(1):47. doi:10.1001/jama.2009.1943

[44] Gentile S. Untreated depression during pregnancy: Short- and long-term effects in offspring. A systematic review. Neuroscience. 2017;342:154–166. doi:10.1016/j.neuroscience.2015.09.001

[45] Chan J, Natekar A, Einarson A, Koren G. Risks of untreated depression in pregnancy. Can Fam Physician. 2014;60(3):242–243.

