## Supplementary materials for "Timing of selective serotonin reuptake inhibitor use and risk for preterm birth and related adverse events"

### Supplemental tables

**Table S1 SNOMED diagnostic codes and RxNorm codes for medication order**

| Diagnosis | SNOMED Code^a^ |
| --- | --- |
| anemia | 271737000 |
| adjustment disorder | 17226007 |
| asthma | 195967001 |
| anxiety disorder | 197480006 |
| bipolar | 13746004 |
| cardiovascular disease | 233604007 |
| chronic lung disease | 413839001 |
| cystic fibrosis | 190905008 |
| depression | 69322001 |
| diabetes | 46635009, 44054006 |
| leukemia | 93143009 |
| pneumonia | 233604007 |
| psychosis | 69322001 |
| renal diseases | 90708001 |
| sepsis | 91302008 |
| sickle cell diseases | 417357006 |

| Medication | RxNorm Code |
| --- | --- |
| N06AA | 17698, 704, 722, 19895, 2597, 3247, 3332, 3634, 3638, 5691, 5979, 6465, 6646, 446248, 7531, 7674, 8886, 35242, 10834 |
| N06AB | citalopram: 2556, escitalopram: 321988, fluoxetine: 4493, fluvoxamine: 42355, paroxetine: 32937, sertraline: 36437 |
| N06AF | 6011, 7394, 8123, 10734 |
| N06AG | 30121, 38382 |
| N06AX | 94, 47111, 42347, 734064, 72625, 2119365, 29434, 6929, 588250, 30031, 15996, 31565, 7500, 60842, 258326, 38252, 10737, 10898, 39786, 1086769, 11196, 1455099 |

**Table S2 Result of contingency table chi-squared test, Welch’s t-test and one-way ANOVA test on distribution of variable across exposure groups and outcome groups**

|  | **Exposure group** | | | | **Outcome group** |
| --- | --- | --- | --- | --- | --- |
|  | **no early-only late (P-value)^c^** | **no**  **late**  **(P-value)^d^** | **no**  **early-only**  **(P-value)^d^** | **early-only**  **late**  **(P-value)^d^** | **non-PTB PTB**  **(P-value)^e^** |
| **pregnancy characteristics** |  |  |  |  |  |
| parity ^a^ | 8.7E-01 | 7.5E-01 | 7.5E-01 | 7.5E-01 | 1.0E-20 |
| parity group ^b^ | 2.7E-01 | 3.1E-01 | 3.1E-01 | 7.6E-01 | 3.2E-38 |
| preterm birth history ^a^ | 6.6E-01 | 9.5E-01 | 5.3E-01 | 5.3E-01 | 4.2E-43 |
| delivery year ^b^ | 4.5E-07 | 2.7E-04 | 1.1E-04 | 8.4E-04 | 7.8E-02 |
| **maternal characteristics** |  |  |  |  |  |
| maternal age ^a^ | 1.3E-17 | 3.6E-13 | 1.3E-02 | 6.6E-12 | 1.4E-01 |
| race group ^b^ | 7.9E-07 | 6.1E-08 | 2.9E-01 | 1.2E-01 | 5.9E-02 |
| ethnic group ^b^ | 6.0E-06 | 6.0E-06 | 9.3E-01 | 1.1E-03 | 5.4E-01 |
| insurance group ^b^ | 5.0E-06 | 1.5E-05 | 7.4E-01 | 2.4E-03 | 9.9E-05 |
| pregravid BMI ^a^ | 3.4E-01 | 4.8E-01 | 9.5E-01 | 5.7E-01 | 5.8E-02 |
| pregravid BMI group ^b^ | 2.2E-01 | 3.0E-01 | 6.5E-01 | 5.5E-01 | 4.3E-03 |
| smoking status ^b^ | 2.2E-01 | 3.0E-01 | 6.5E-01 | 5.5E-01 | 4.3E-03 |
| illegal drug use status ^b^ | 7.6E-01 | 7.6E-01 | 7.6E-01 | 7.6E-01 | 9.0E-02 |
| ethanol consumption status ^b^ | 2.3E-11 | 5.0E-09 | 1.4E-05 | 1.4E-01 | 6.8E-01 |
| social vulnerability index ^b^ | 2.8E-5 | 5.9E-05 | 7.5E-01 | 3.7E-03 | 7.2E-01 |
| **comorbidities** |  |  |  |  |  |
| anemia ^b^ | 6.4E-02 | 6.4E-02 | 5.7E-01 | 6.3E-01 | 5.3E-01 |
| asthma ^b^ | 8.0E-01 | 9.6E-01 | 8.7E-01 | 8.7E-01 | 3.3E-01 |
| cardiovascular diseases ^b^ | 4.4E-01 | 6.5E-01 | 8.5E-01 | 8.5E-01 | 3.3E-05 |
| cystic fibrosis ^b^ | 3.5E-01 | 5.6E-01 | 8.0E-01 | N/A | 4.6E-01 |
| diabetes ^b^ | 4.3E-03 | 2.9E-02 | 1.9E-01 | 2.9E-02 | 2.0E-28 |
| leukemia ^b^ | N/A | N/A | N/A | N/A | N/A |
| pneumonia ^b^ | 5.1E-01 | 6.1E-01 | 5.1E-01 | 5.1E-01 | 6.3E-02 |
| renal diseases ^b^ | 1.5E-01 | 1.6E-01 | 5.1E-01 | 7.8E-01 | 7.7E-02 |
| sepsis ^b^ | 2.0E-01 | 2.0E-01 | 6.3E-01 | 6.3E-01 | 5.8E-01 |
| sickle cell diseases ^b^ | 4.8E-01 | 9.2E-01 | N/A | 4.8E-01 | 1.7E-01 |
| **pre-pregnancy condition** |  |  |  |  |  |
| adjustment disorder ^b^ | 3.5E-01 | 9.2E-01 | 3.3E-01 | 3.3E-01 | 7.7E-01 |
| anxiety disorder ^b^ | 2.1E-10 | 9.3E-11 | 3.5E-02 | 6.7E-02 | 3.1E-01 |
| PHQ-9 score ^b^ | 2.4E-02 | 8.2E-01 | 2.3E-02 | 2.3E-02 | 7.9E-01 |
| PHQ-9 category ^b^ | 3.6E-01 | 8.0E-01 | 2.7E-01 | 2.7E-01 | 7.5E-01 |
| N06AA exposure ^b^ | 2.9E-07 | 3.2E-07 | 8.2E-02 | 2.1E-01 | 1.1E-01 |
| N06AB exposure ^b^ | 0.0E+00 | 0.0E+00 | 0.0E+00 | 1.3E-18 | 9.3E-01 |
| N06AG exposure ^b^ | N/A | N/A | N/A | N/A | N/A |
| N06AF exposure ^b^ | N/A | N/A | N/A | N/A | N/A |
| N06AX exposure ^b^ | 4.4E-16 | 5.0E-12 | 5.0E-09 | 8.1E-02 | 1.9E-01 |
| **prenatal depression** |  |  |  |  |  |
| late pregnancy depression ^b^ | 6.2E-51 | 3.1E-51 | 3.8E-07 | 1.0E-05 | 3.9E-02 |
| PHQ-4 score ^a^ | 1.4E-03 | 6.1E-03 | 3.4E-01 | 6.1E-03 | 2.0E-01 |
| PHQ-4 category ^b^ | 3.2E-02 | 6.3E-02 | 7.1E-01 | 9.0E-02 | 3.0E-01 |

**Table S3 Result of the main analysis on preterm birth, gestational age at birth (top), low birthweight and small for gestational age (bottom)**

| reference group (n) | comparison group (n) | unadjusted PTB OR | adjusted PTB OR | adjusted PTB OR  p-value | unadjusted GA β | adjusted GA β | adjusted GA β p-value |
| --- | --- | --- | --- | --- | --- | --- | --- |
| no SSRI exposure (3122) | early-only SSRI exposure (690) | 0.8 [0.5,1.0]  + | 1.4 [0.8,2.4] | 2.3E-01 | 1.1 [0.0,2.1] * | -0.2 [-1.8,1.5] | 8.3E-01 |
| no-late SSRI exposure (3812) | late SSRI exposure (2596) | 1.4 [1.2,1.7] **** | 1.6 [1.3,1.9] **** | 3.6E-05 | -1.9 [-2.6,-1.3] ***** | -2.3 [-3.1,-1.5] ***** | 1.3E-08 |
| no SSRI exposure (3122) | late SSRI exposure (2596) | 1.4 [1.1,1.6] *** | 1.7 [1.3,2.2] **** | 1.2E-05 | -1.8 [-2.4,-1.1] ***** | -2.6 [-3.6,-1.5] ***** | 7.7E-07 |
| early-only SSRI exposure (690) | late SSRI exposure (2596) | 1.8 [1.3,2.5] *** | 1.5 [1.1,2.2] ** | 1.2E-02 | -2.8 [-3.9,-1.7] ***** | -1.9 [-3.1,-0.8] *** | 7.2E-04 |
| no SSRI exposure (3122) | any SSRI exposure (3286) | 1.2 [1.0,1.5] * | 1.6 [1.2,2.0] *** | 4.0E-04 | -1.2 [-1.8,-0.6] *** | -2.3 [-3.2,-1.3] ***** | 6.2E-06 |

| reference group (n) | comparison group (n) | unadjusted LBW OR | adjusted LBW OR | adjusted LBW OR  p-value | unadjusted SGA OR | adjusted SGA OR | unadjusted SGA OR p-value |
| --- | --- | --- | --- | --- | --- | --- | --- |
| no SSRI exposure (3122) | early-only SSRI exposure (690) | 0.7 [0.4,1.0]+ | 1.0 [0.5,2.0] | 9.6E-01 | 0.8 [0.6,1.0]+ | 1.0 [0.6,1.5] | 9.7E-01 |
| no-late SSRI exposure (3812) | late SSRI exposure (2596) | 1.3 [1.1,1.6] ** | 1.7 [1.3,2.2] *** | 1.0E-04 | 1.0 [0.9,1.2] | 1.2 [1.0,1.5]+ | 6.6E-02 |
| no SSRI exposure (3122) | late SSRI exposure (2596) | 1.3 [1.0,1.6] * | 1.7 [1.3,2.4] *** | 6.8E-04 | 1.0 [0.8,1.1] | 1.2 [0.9,1.5] | 2.1E-01 |
| early-only SSRI exposure (690) | late SSRI exposure (2596) | 1.9 [1.2,2.9] ** | 1.6 [1.1,2.5] * | 2.3E-02 | 1.3 [1.0,1.7] | 1.2 [0.9,1.6] | 2.0E-01 |
| no SSRI exposure (3122) | any SSRI exposure (3286) | 1.1 [0.9,1.4] | 1.6 [1.2,2.2] ** | 1.9E-03 | 0.9 [0.8,1.1] | 1.2 [0.9,1.5] | 2.1E-01 |

**Table S4 Result of the exposure group supplementary analysis on preterm birth, gestational age at birth (top), low birthweight and small for gestational age (bottom**)

| reference group (n) | comparison group (n) | unadjusted PTB OR | adjusted PTB OR | adjusted PTB OR  p-value | unadjusted GA β | adjusted GA β | adjusted GA β p-value |
| --- | --- | --- | --- | --- | --- | --- | --- |
| no SSRI exposure (3122) | both SSRI exposure  (2259) | 1.3 [1.1,1.6] ** | 1.8 [1.3,2.5] *** | 9.1E-04 | -1.5 [-2.2,-0.8] **** | -2.2 [-3.5,-1.0] *** | 4.3E-04 |
| no SSRI exposure (3122) | late-only SSRI exposure  (337) | 1.7 [1.2,2.3] ** | 1.6 [1.1,2.2] ** | 7.7E-03 | -3.5 [-5.0,-2.0] ***** | -3.1 [-4.6,-1.6] **** | 3.9E-05 |
| both SSRI exposure (2259) | late-only SSRI exposure  (337) | 1.3 [0.9,1.8] | 1.1 [0.7,1.8] | 6.6E-01 | -2.0 [-3.5,-0.5] ** | -0.3 [-2.5,1.8] | 7.6E-01 |
| early-only SSRI exposure  (690) | late-only SSRI exposure  (337) | 2.2 [1.4,3.4] *** | 1.9 [0.9,4.2]+ | 9.5E-02 | -4.6 [-6.3,-2.8] ***** | -1.9 [-5.1,1.3] | 2.4E-01 |
| early-only SSRI exposure  (690) | both SSRI exposure (2252) | 1.7 [1.3,2.4] *** | 1.5 [1.1,2.2] * | 1.1E-02 | -2.5 [-3.6,-1.5] ***** | -1.9 [-3.0,-0.9] *** | 4.4E-04 |

| reference group (n) | comparison group (n) | unadjusted LBW OR | adjusted LBW OR | adjusted LBW OR  p-value | unadjusted SGA β | adjusted SGA β | adjusted SGA β p-value |
| --- | --- | --- | --- | --- | --- | --- | --- |
| no SSRI exposure (3122) | both SSRI exposure  (2259) | 1.2 [1.0,1.5] | 1.7 [1.2,2.6] ** | 8.5E-03 | 0.9 [0.8,1.1] | 1.1 [0.8,1.5] | 4.1E-01 |
| no SSRI exposure (3122) | late-only SSRI exposure  (337) | 1.6 [1.1,2.5] * | 1.6 [1.1,2.5] * | 2.1E-02 | 1.1 [0.8,1.6] | 1.2 [0.8,1.6] | 3.6E-01 |
| both SSRI exposure (2259) | late-only SSRI exposure  (337) | 1.4 [0.9,2.1] | 0.9 [0.5,1.7] | 7.8E-01 | 1.2 [0.8,1.7] | 0.9 [0.6,1.5] | 7.2E-01 |
| early-only SSRI exposure  (690) | late-only SSRI exposure  (337) | 2.5 [1.4,4.2] ** | 1.2 [0.4,3.5] | 7.1E-01 | 1.5 [1.0,2.2]+ | 0.8 [0.3,1.7] | 5.0E-01 |
| early-only SSRI exposure  (690) | both SSRI exposure (2252) | 1.8 [1.2,2.7] ** | 1.7 [1.1,2.6] * | 2.0E-02 | 1.2 [0.9,1.6] | 1.2 [0.9,1.6] | 1.8E-01 |

**Table S5 Result of drug specific analyses**

| citalopram | | | | | | | |
| --- | --- | --- | --- | --- | --- | --- | --- |
| reference group (n) | comparison group (n) | unadjusted PTB OR | adjusted PTB OR | adjusted PTB OR  p-value | unadjusted GA β | adjusted GA β | adjusted GA β p-value |
| no citalopram exposure (3122) | early-only citalopram exposure (196) | 1.1 [0.7,1.9] | 2.1 [1.0,4.1] * | 4.2E-02 | 1.0 [-0.8,2.8] | 0.3 [-2.1,2.6] | 8.3E-01 |
| no-late citalopram exposure (3318) | late citalopram exposure (436) | 0.9 [0.7,1.4] | 1.1 [0.7,1.7] | 6.9E-01 | -0.7 [-1.9,0.6] | -0.9 [-2.5,0.6] | 2.3E-01 |
| no citalopram exposure (3122) | late citalopram exposure (436) | 1.0 [0.7,1.4] | 1.3 [0.8,2.2] | 3.1E-01 | -0.6 [-1.9,0.6] | -0.9 [-2.6,0.8] | 3.0E-01 |
| early-only citalopram exposure (196) | late citalopram exposure (436) | 0.8 [0.5,1.5] | 0.7 [0.4,1.4] | 3.2E-01 | -1.6 [-3.6,0.3] | -1.0 [-3.1,1.2] | 3.7E-01 |
| no citalopram exposure (3122) | any citalopram exposure (632) | 1.0 [0.7,1.4] | 1.4 [0.9,2.3] | 1.4E-01 | -0.1 [-1.2,0.9] | -0.6 [-2.1,1.0] | 4.9E-01 |

| citalopram (continued) | | | | | | | |
| --- | --- | --- | --- | --- | --- | --- | --- |
| reference group (n) | comparison group (n) | unadjusted LBW OR | adjusted LBW OR | adjusted SGA OR  p-value | unadjusted SGA OR | adjusted SGA OR | adjusted SGA OR  p-value |
| no citalopram exposure (3122) | early-only citalopram exposure (196) | 0.5 [0.2,1.2] | 0.8 [0.3,2.2] | 6.5E-01 | 0.7 [0.4,1.2] | 0.8 [0.4,1.6] | 5.6E-01 |
| no-late citalopram exposure (3318) | late citalopram exposure (436) | 1.4 [0.9,2.0] | 1.8 [1.1,3.1] * | 2.2E-02 | 1.2 [0.9,1.5] | 1.5 [1.0,2.2] * | 3.7E-02 |
| no citalopram exposure (3122) | late citalopram exposure (436) | 1.3 [0.9,2.0] | 1.7 [0.9,2.9]+ | 8.1E-02 | 1.1 [0.8,1.5] | 1.4 [0.9,2.1] | 1.1E-01 |
| early-only citalopram exposure (196) | late citalopram exposure (436) | 2.4 [1.0,6.0] * | 2.3 [0.9,5.9]+ | 8.1E-02 | 1.5 [0.9,2.7] | 1.4 [0.8,2.6] | 2.4E-01 |
| no citalopram exposure (3122) | any citalopram exposure (632) | 1.1 [0.8,1.6] | 1.4 [0.8,2.5] | 2.1E-01 | 1.0 [0.8,1.3] | 1.3 [0.9,1.9] | 2.0E-01 |

| escitalopram | | | | | | | |
| --- | --- | --- | --- | --- | --- | --- | --- |
| reference group (n) | comparison group (n) | unadjusted PTB OR | adjusted PTB OR | adjusted PTB OR  p-value | unadjusted GA β | adjusted GA β | adjusted GA β p-value |
| no escitalopram exposure (3122) | early-only escitalopram exposure (124) | 1.0 [0.5,1.9] | 2.6 [1.1,6.2] * | 2.7E-02 | -0.4 [-2.6,1.9] | -2.1 [-4.9,0.7] | 1.1E-01 |
| no-late escitalopram exposure (3246) | late escitalopram exposure (319) | 1.1 [0.8,1.7] | 1.4 [0.8,2.3] | 1.9E-01 | -1.0 [-2.4,0.4] | -1.5 [-3.3,0.3] | 1.0E-01 |
| no escitalopram exposure (3122) | late escitalopram exposure (319) | 1.1 [0.8,1.7] | 1.5 [0.9,2.7] | 1.2E-01 | -1.0 [-2.5,0.4] | -1.9 [-3.8,0.0]  + | 5.8E-02 |
| early-only escitalopram exposure (124) | late escitalopram exposure (319) | 1.1 [0.5,2.3] | 0.9 [0.4,1.9] | 7.3E-01 | -0.6 [-2.9,1.6] | 0.2 [-2.2,2.6] | 8.6E-01 |
| no escitalopram exposure (3122) | any escitalopram exposure (443) | 1.1 [0.8,1.6] | 1.8 [1.1,2.9] * | 2.8E-02 | -0.8 [-2.1,0.4] | -2.0 [-3.8,-0.3] * | 2.5E-02 |

| escitalopram (continued) | | | | | | | |
| --- | --- | --- | --- | --- | --- | --- | --- |
| reference group (n) | comparison group (n) | unadjusted LBW OR | adjusted LBW OR | adjusted LBW OR  p-value | unadjusted SGA OR | adjusted  SGA OR | adjusted SGA OR p-value |
| no escitalopram exposure (3122) | early-only escitalopram exposure (124) | 0.7 [0.3,1.8] | 1.3 [0.4,4.2] | 6.1E-01 | 0.8 [0.4,1.4] | 0.9 [0.4,1.9] | 7.9E-01 |
| no-late escitalopram exposure (3246) | late escitalopram exposure (319) | 1.1 [0.7,1.8] | 1.9 [1.0,3.6] * | 3.6E-02 | 1.1 [0.8,1.5] | 1.4 [0.9,2.1] | 1.7E-01 |
| no-escitalopram exposure (3122) | late escitalopram exposure (319) | 1.1 [0.7,1.8] | 2.0 [1.0,3.9] * | 3.6E-02 | 1.1 [0.8,1.5] | 1.4 [0.9,2.2] | 1.8E-01 |
| early-only escitalopram exposure (124) | late escitalopram exposure (319) | 1.6 [0.6,4.3] | 1.4 [0.5,4.0] | 5.6E-01 | 1.4 [0.7,2.8] | 1.4 [0.7,3.0] | 3.2E-01 |
| no escitalopram exposure (3122) | any escitalopram exposure (443) | 1.0 [0.7,1.6] | 1.9 [1.0,3.6] * | 4.2E-02 | 1.0 [0.7,1.4] | 1.2 [0.8,1.9] | 4.2E-01 |

| fluoxetine | | | | | | | |
| --- | --- | --- | --- | --- | --- | --- | --- |
| reference group (n) | comparison group (n) | unadjusted PTB OR | adjusted PTB OR | adjusted PTB OR  p-value | unadjusted GA β | adjusted  GA β | adjusted GA β p-value |
| no fluoxetine exposure (3122) | early-only fluoxetine exposure (164) | 0.8 [0.4,1.4] | 1.5 [0.7,3.5] | 3.0E-01 | 0.4 [-1.6,2.3] | -1.1 [-3.6,1.4] | 3.9E-01 |
| no fluoxetine exposure (3286) | late fluoxetine exposure (482) | 1.5 [1.1,2.0] ** | 1.9 [1.3,2.8] ** | 1.1E-03 | -2.6 [-3.9,-1.4] **** | -3.2 [-4.7,-1.7] **** | 4.1E-05 |
| early-only fluoxetine exposure (3122) | late fluoxetine exposure (482) | 1.5 [1.1,2.0] * | 2.0 [1.3,3.1] ** | 1.2E-03 | -2.6 [-3.8,-1.4] **** | -3.4 [-5.1,-1.7] **** | 6.7E-05 |
| early-only fluoxetine exposure (164) | late fluoxetine exposure (482) | 1.9 [1.0,3.8]+ | 1.7 [0.9,3.4] | 1.3E-01 | -3.0 [-5.5,-0.4] * | -2.3 [-5.0,0.3]+ | 8.6E-02 |
| no fluoxetine exposure (3122) | any fluoxetine exposure (646) | 1.3 [1.0,1.7]  + | 1.9 [1.3,2.9] ** | 2.3E-03 | -1.8 [-2.9,-0.7] *** | -3.0 [-4.6,-1.4] *** | 2.3E-04 |

| fluoxetine (continued) | | | | | | | |
| --- | --- | --- | --- | --- | --- | --- | --- |
| reference group (n) | comparison group (n) | unadjusted LBW OR | adjusted LBW OR | adjusted LBW OR p-value | unadjusted SGA OR | adjusted SGA OR | adjusted SGA OR p-value |
| no fluoxetine exposure (3122) | early-only fluoxetine exposure (164) | 0.7 [0.3,1.6] | 1.0 [0.4,2.6] | 9.7E-01 | 0.9 [0.5,1.4] | 1.0 [0.5,1.8] | 9.4E-01 |
| no fluoxetine exposure (3286) | late fluoxetine exposure (482) | 1.1 [0.8,1.7] | 1.6 [0.9,2.6]+ | 8.7E-02 | 0.9 [0.7,1.2] | 1.0 [0.7,1.5] | 8.5E-01 |
| early-only fluoxetine exposure (3122) | late fluoxetine exposure (482) | 1.1 [0.7,1.7] | 1.7 [1.0,2.9]+ | 7.4E-02 | 0.9 [0.6,1.2] | 1.0 [0.7,1.5] | 9.3E-01 |
| early-only fluoxetine exposure (164) | late fluoxetine exposure (482) | 1.5 [0.6,3.5] | 1.3 [0.5,3.0] | 6.0E-01 | 1.0 [0.6,1.8] | 1.1 [0.6,2.1] | 6.7E-01 |
| no fluoxetine exposure (3122) | any fluoxetine exposure (646) | 1.0 [0.7,1.5] | 1.6 [0.9,2.7]+ | 9.1E-02 | 0.9 [0.7,1.2] | 1.0 [0.7,1.5] | 8.7E-01 |

| paroxetine | | | | | | | |
| --- | --- | --- | --- | --- | --- | --- | --- |
| reference group (n) | comparison group (n) | unadjusted PTB OR | adjusted PTB OR | adjusted PTB OR p-value | unadjusted GA β | adjusted GA β | adjusted GA β p-value |
| no paroxetine exposure (3122) | early-only paroxetine exposure (54) | 1.3 [0.6,3.1] | 2.4 [0.9,6.5]+ | 9.3E-02 | 0.4 [-3.0,3.8] | -0.9 [-4.7,2.9] | 6.4E-01 |
| no-late paroxetine exposure (3176) | late paroxetine exposure (54) | 0.4 [0.1,1.7] | 0.5 [0.1,2.5] | 4.1E-01 | -0.3 [-3.7,3.1] | 0.1 [-4.1,4.3] | 9.7E-01 |
| no paroxetine exposure (3122) | late paroxetine exposure (54) | 0.4 [0.1,1.7] | 0.9 [0.2,4.4] | 8.6E-01 | -0.3 [-3.6,3.1] | -0.9 [-5.2,3.4] | 6.8E-01 |
| early-only paroxetine exposure (54) | late paroxetine exposure (54) | 0.3 [0.1,1.6] | 0.3 [0.0,2.7] | 2.7E-01 | -0.6 [-5.2,3.9] | 1.7 [-4.7,8.2] | 6.0E-01 |
| no paroxetine exposure (3122) | late paroxetine exposure (108) | 0.8 [0.4,1.8] | 1.4 [0.6,3.5] | 4.5E-01 | 0.1 [-2.4,2.5] | -0.6 [-3.6,2.4] | 6.8E-01 |

| paroxetine (continued) | | | | | | | |
| --- | --- | --- | --- | --- | --- | --- | --- |
| reference group (n) | comparison group (n) | unadjusted LBW OR | adjusted LBW OR | adjusted LBW OR p-value | unadjusted SGA OR | adjusted SGA OR | adjusted SGA OR  p-value |
| no paroxetine exposure (3122) | early-only paroxetine exposure (54) | 1.1 [0.3,3.6] | 1.6 [0.4,6.2] | 4.6E-01 | 1.2 [0.5,2.7] | 1.2 [0.5,3.0] | 6.8E-01 |
| no-late paroxetine exposure (3176) | late paroxetine exposure (54) | 1.7 [0.7,4.4] | 2.3 [0.7,7.8] | 1.9E-01 | 1.6 [0.8,3.3] | 1.9 [0.8,4.8] | 1.5E-01 |
| no paroxetine exposure (3122) | late paroxetine exposure (54) | 1.7 [0.7,4.4] | 2.3 [0.7,8.2] | 1.9E-01 | 1.7 [0.8,3.3] | 1.7 [0.7,4.3] | 2.7E-01 |
| early-only paroxetine exposure (54) | late paroxetine exposure (54) | 1.6 [0.4,6.9] | 1.5 [0.2,12.4] | 7.1E-01 | 1.4 [0.5,3.9] | 2.9 [0.6,14.0] | 1.8E-01 |
| no paroxetine exposure (3122) | late paroxetine exposure (108) | 1.4 [0.7,3.0] | 1.9 [0.7,4.9] | 1.9E-01 | 1.4 [0.8,2.5] | 1.4 [0.7,2.8] | 2.9E-01 |

| sertraline | | | | | | | |
| --- | --- | --- | --- | --- | --- | --- | --- |
| reference group (n) | comparison group (n) | unadjusted PTB OR | adjusted PTB OR | adjusted PTB OR p-value | unadjusted GA β | adjusted GA β | adjusted GA β  p-value |
| no sertraline exposure (3122) | early-only sertraline exposure (305) | 0.5 [0.3,0.8] ** | 1.0 [0.5,2.2] | 9.6E-01 | 1.4 [-0.1,2.9]+ | -0.3 [-2.4,1.8] | 7.9E-01 |
| no-late sertraline exposure (3427) | late sertraline exposure (1430) | 1.6 [1.3,1.9] ***** | 1.8 [1.4,2.3] ***** | 5.0E-06 | -2.1 [-2.9,-1.3] ***** | -2.4 [-3.4,-1.4] ***** | 2.0E-06 |
| no sertraline exposure (3122) | late sertraline exposure (1430) | 1.5 [1.2,1.8] **** | 1.8 [1.3,2.4] **** | 7.2E-05 | -2.0 [-2.8,-1.2] ***** | -2.5 [-3.7,-1.4] **** | 1.8E-05 |
| early-only sertraline exposure (305) | late sertraline exposure (1430) | 3.2 [1.8,5.7] **** | 2.5 [1.4,4.4] ** | 2.8E-03 | -3.4 [-5.0,-1.8] **** | -2.2 [-3.9,-0.5] * | 1.1E-02 |
| no  sertraline exposure (3122) | any sertraline exposure (1735) | 1.3 [1.1,1.6] ** | 1.7 [1.2,2.2] *** | 5.2E-04 | -1.4 [-2.2,-0.6] *** | -2.2 [-3.3,-1.1] *** | 1.1E-04 |

| sertraline (continued) | | | | | | | |
| --- | --- | --- | --- | --- | --- | --- | --- |
| reference group (n) | comparison group (n) | unadjusted LBW OR | adjusted LBW OR | adjusted LBW OR p-value | unadjusted SGA OR | adjusted SGA OR | adjusted SGA OR p-value |
| no sertraline exposure (3122) | early-only sertraline exposure (305) | 0.7 [0.4,1.3] | 1.0 [0.4,2.3] | 9.7E-01 | 0.6 [0.4,1.0] * | 0.9 [0.5,1.5] | 6.1E-01 |
| no-late sertraline exposure (3427) | late sertraline exposure (1430) | 1.4 [1.1,1.8] * | 1.8 [1.3,2.5] *** | 1.7E-04 | 1.0 [0.8,1.2] | 1.2 [1.0,1.6]+ | 7.3E-02 |
| no sertraline exposure (3122) | late sertraline exposure (1430) | 1.3 [1.0,1.7] * | 1.9 [1.4,2.7] *** | 2.1E-04 | 0.9 [0.8,1.1] | 1.2 [0.9,1.6] | 2.3E-01 |
| early-only sertraline exposure (305) | late sertraline exposure (1430) | 1.9 [1.1,3.6] * | 1.7 [0.9,3.2] | 1.1E-01 | 1.5 [0.9,2.3]+ | 1.4 [0.9,2.2] | 1.6E-01 |
| no  sertraline exposure (3122) | any sertraline exposure (1735) | 1.2 [1.0,1.6]  + | 1.9 [1.3,2.6] *** | 5.5E-04 | 0.9 [0.7,1.1] | 1.2 [0.9,1.5] | 2.4E-01 |

**Table S6** Result of sensitivity analyses

| subsample of women with depression diagnosis during late pregnancy^a^ | | | | | | | |
| --- | --- | --- | --- | --- | --- | --- | --- |
| reference group (n) | comparison group (n) | unadjusted PTB OR | adjusted PTB OR | adjusted PTB OR p-value | unadjusted GA β | adjusted GA β | adjusted GA β  p-value |
| no-late SSRI exposure (2057) | late SSRI exposure (1862) | 1.2 [1.0,1.5]+ | 1.3 [1.0,1.6]+ | 7.9E-02 | -1.5 [-2.3,-0.7] *** | -1.7 [-2.7,-0.7] *** | 8.1E-04 |
| no SSRI exposure (1622) | late SSRI exposure (1862) | 1.2 [0.9,1.4] | 1.3 [1.0,1.8]+ | 8.5E-02 | -1.2 [-2.1,-0.4] ** | -1.8 [-3.1,-0.5] ** | 6.2E-03 |
| early-only SSRI exposure (435) | late SSRI exposure (1862) | 1.5 [1.0,2.2] * | 1.3 [0.9,1.9] | 2.1E-01 | -2.4 [-3.7,-1.0] *** | -1.6 [-3.0,-0.2] * | 2.2E-02 |

| subsample of women with depression diagnosis during late pregnancy^a^ (continued) | | | | | | | |
| --- | --- | --- | --- | --- | --- | --- | --- |
| reference group (n) | comparison group (n) | unadjusted LBW OR | adjusted LBW OR | adjusted LBW OR p-value | unadjusted SGA OR | adjusted SGA OR | adjusted SGA OR p-value |
| no-late SSRI exposure (2057) | late SSRI exposure (1862) | 1.2 [0.9,1.5] | 1.4 [1.0,1.9] * | 4.7E-02 | 0.9 [0.7,1.1] | 1.0 [0.8,1.3] | 7.5E-01 |
| no SSRI exposure (1622) | late SSRI exposure (1862) | 1.1 [0.9,1.5] | 1.4 [1.0,2.1]+ | 6.1E-02 | 0.9 [0.7,1.1] | 1.0 [0.8,1.4] | 9.0E-01 |
| early-only SSRI exposure (435) | late SSRI exposure (1862) | 1.5 [0.9,2.4] | 1.3 [0.8,2.2] | 2.6E-01 | 1.1 [0.8,1.5] | 1.0 [0.7,1.5] | 8.4E-01 |

| subsample of women with phq-9 score during two-year prepregnancy period^b^ | | | | | | | |
| --- | --- | --- | --- | --- | --- | --- | --- |
| reference group (n) | comparison group (n) | unadjusted PTB OR | adjusted PTB OR | adjusted PTB OR p-value | unadjusted GA β | adjusted GA β | adjusted GA β  p-value |
| no SSRI exposure (836) | early-only SSRI exposure (290) | 0.9 [0.5,1.5] | 1.4 [0.6,3.7] | 4.6E-01 | 0.5 [-1.1,2.1] | -0.8 [-3.7,2.1] | 5.9E-01 |
| no-late SSRI exposure (1126) | late SSRI exposure (917) | 1.4 [1.0,1.8]  + | 1.5 [1.0,2.1] * | 4.9E-02 | -2.3 [-3.3,-1.2] ***** | -2.6 [-3.9,-1.3] ***** | 9.8E-05 |
| no SSRI exposure (836) | late SSRI exposure (917) | 1.3 [0.9,1.8] | 1.7 [1.0,2.7] * | 4.0E-02 | -2.1 [-3.3,-1.0] **** | -3.2 [-5.0,-1.5] **** | 3.4E-04 |
| early-only SSRI exposure (290) | late SSRI exposure (917) | 1.5 [0.9,2.4] | 1.3 [0.8,2.1] | 3.5E-01 | -2.6 [-4.3,-0.9] *** | -1.8 [-3.6,-0.0] * | 4.5E-02 |
| no SSRI exposure (836) | any SSRI exposure (1207) | 1.2 [0.9,1.6] | 1.6 [1.0,2.5]  + | 7.4E-02 | -1.5 [-2.6,-0.4] ** | -2.8 [-4.6,-1.1] *** | 1.4E-03 |

| subsample of women with phq-9 score during two year prepregnancy period^b^ (continued) | | | | | | | |
| --- | --- | --- | --- | --- | --- | --- | --- |
| reference group (n) | comparison group (n) | unadjusted LBW OR | adjusted LBW OR | adjusted LBW OR  p-value | unadjusted SGA OR | adjusted SGA OR | adjusted SGA OR  p-value |
| no SSRI exposure (836) | early-only SSRI exposure (290) | 0.9 [0.5,1.6] | 2.0 [0.6,6.0] | 2.4E-01 | 0.8 [0.5,1.3] | 0.7 [0.3,1.5] | 3.7E-01 |
| no-late SSRI exposure (1126) | late SSRI exposure (917) | 1.2 [0.8,1.7] | 1.5 [1.0,2.4]  + | 6.2E-02 | 1.2 [0.9,1.5] | 1.3 [0.9,1.8] | 1.6E-01 |
| no SSRI exposure (836) | late SSRI exposure (917) | 1.1 [0.8,1.6] | 1.8 [1.0,3.2] * | 4.7E-02 | 1.1 [0.8,1.5] | 1.1 [0.7,1.8] | 6.0E-01 |
| early-only SSRI exposure (290) | late SSRI exposure (917) | 1.3 [0.7,2.3] | 1.2 [0.7,2.2] | 5.5E-01 | 1.3 [0.9,2.0] | 1.4 [0.9,2.2] | 1.4E-01 |
| no SSRI exposure (836) | any SSRI exposure (1207) | 1.1 [0.7,1.5] | 1.8 [1.0,3.2] * | 3.8E-02 | 1.0 [0.8,1.4] | 1.1 [0.7,1.7] | 7.7E-01 |

| subsample of women with phq-4 score during pregnancy^c^ | | | | | | | |
| --- | --- | --- | --- | --- | --- | --- | --- |
| reference group (n) | comparison group (n) | unadjusted PTB OR | adjusted PTB OR | adjusted PTB OR p-value | unadjusted GA β | adjusted GA β | adjusted GA β  p-value |
| no SSRI exposure (1238) | early-only SSRI exposure (245) | 0.8 [0.5,1.2] | 1.1 [0.5,2.4] | 8.0E-01 | 1.3 [-0.6,3.2] | 0.6 [-2.6,3.8] | 7.3E-01 |
| no-late SSRI exposure (1483) | late SSRI exposure (1007) | 1.2 [0.9,1.5] | 1.3 [0.9,1.7] | 1.2E-01 | -2.1 [-3.3,-0.9] *** | -2.5 [-3.9,-1.0] *** | 7.4E-04 |
| no SSRI exposure (1238) | late SSRI exposure (1007) | 1.2 [0.9,1.5] | 1.3 [0.9,1.8] | 1.9E-01 | -1.9 [-3.1,-0.7] ** | -2.8 [-4.6,-1.0] ** | 2.5E-03 |
| early-only SSRI exposure (245) | late SSRI exposure (1007) | 1.5 [0.9,2.4]+ | 1.3 [0.8,2.2] | 2.8E-01 | -3.2 [-5.3,-1.1] ** | -2.2 [-4.4,0.0]+ | 5.1E-02 |
| no SSRI exposure (1238) | any SSRI exposure (1252) | 1.1 [0.8,1.4] | 1.2 [0.9,1.8] | 2.3E-01 | -1.3 [-2.4,-0.1] * | -2.4 [-4.1,-0.6] ** | 7.4E-03 |

| subsample of women with phq-4 score during pregnancy^c^ (continued) | | | | | | | |
| --- | --- | --- | --- | --- | --- | --- | --- |
| reference group (n) | comparison group (n) | unadjusted LBW OR | adjusted LBW OR | adjusted LBW OR p-value | unadjusted SGA OR | adjusted SGA OR | adjusted SGA OR p-value |
| no SSRI exposure (1238) | early-only SSRI exposure (245) | 0.5 [0.3,1.0]+ | 0.6 [0.2,1.6] | 3.2E-01 | 0.4 [0.3,0.7] ** | 0.6 [0.3,1.4] | 2.4E-01 |
| no-late SSRI exposure (1483) | late SSRI exposure (1007) | 1.3 [1.0,1.8] * | 1.7 [1.2,2.4] ** | 6.4E-03 | 1.0 [0.8,1.3] | 1.3 [0.9,1.7] | 1.2E-01 |
| no SSRI exposure (1238) | late SSRI exposure (1007) | 1.2 [0.9,1.7] | 1.6 [1.1,2.5] * | 2.4E-02 | 0.9 [0.7,1.2] | 1.1 [0.7,1.5] | 7.4E-01 |
| early-only SSRI exposure (245) | late SSRI exposure (1007) | 2.3 [1.2,4.5] * | 1.9 [0.9,3.8]+ | 7.0E-02 | 2.0 [1.2,3.5] ** | 1.9 [1.1,3.3] * | 1.8E-02 |
| no SSRI exposure (1238) | any SSRI exposure (1252) | 1.1 [0.8,1.5] | 1.5 [1.0,2.3]+ | 5.1E-02 | 0.8 [0.6,1.0]+ | 1.0 [0.7,1.4] | 9.9E-01 |

| subsample of women exposed to SSRI only^d^ | | | | | | | |
| --- | --- | --- | --- | --- | --- | --- | --- |
| reference group (n) | comparison group (n) | unadjusted PTB OR | adjusted PTB OR | adjusted PTB OR p-value | unadjusted GA β | adjusted GA β | adjusted GA β  p-value |
| no SSRI exposure (2350) | early-only SSRI exposure (582) | 0.9 [0.6,1.3] | 1.8 [0.9,3.6]+ | 8.0E-02 | 0.5 [-0.6,1.6] | -0.6 [-2.4,1.3] | 5.6E-01 |
| no-late SSRI exposure (2932) | late SSRI exposure (2133) | 1.5 [1.2,1.8] **** | 1.6 [1.2,2.0] *** | 3.3E-04 | -2.1 [-2.8,-1.4] ***** | -2.3 [-3.2,-1.4] ***** | 1.8E-07 |
| no SSRI exposure (2350) | late SSRI exposure (2133) | 1.5 [1.2,1.8] *** | 1.9 [1.4,2.5] *** | 1.3E-04 | -2.0 [-2.8,-1.3] ***** | -3.0 [-4.1,-1.8] ***** | 6.1E-07 |
| early-only SSRI exposure (582) | late SSRI exposure (2133) | 1.6 [1.1,2.3] ** | 1.3 [0.9,1.9] | 1.1E-01 | -2.5 [-3.7,-1.4] **** | -1.6 [-2.7,-0.4] ** | 9.4E-03 |
| no SSRI exposure (2350) | any SSRI exposure (2715) | 1.3 [1.1,1.6] ** | 1.8 [1.3,2.5] *** | 1.5E-04 | -1.5 [-2.2,-0.8] **** | -2.7 [-3.9,-1.6] ***** | 1.9E-06 |

| subsample of women exposed to SSRI only^d^ (continued) | | | | | | | |
| --- | --- | --- | --- | --- | --- | --- | --- |
| reference group (n) | comparison group (n) | unadjusted LBW OR | adjusted LBW OR | adjusted PTB OR p-value | unadjusted SGA OR | adjusted SGA OR | adjusted SGA OR p-value |
| no SSRI exposure (2350) | early-only SSRI exposure (582) | 0.9 [0.6,1.4] | 1.4 [0.6,3.0] | 4.6E-01 | 0.8 [0.6,1.1] | 0.9 [0.5,1.5] | 6.5E-01 |
| no-late SSRI exposure (2932) | late SSRI exposure (2133) | 1.3 [1.1,1.7] * | 1.6 [1.2,2.2] ** | 1.9E-03 | 1.0 [0.8,1.2] | 1.2 [0.9,1.5] | 1.7E-01 |
| no SSRI exposure (2350) | late SSRI exposure (2133) | 1.3 [1.0,1.7] * | 1.9 [1.3,2.8] *** | 9.4E-04 | 1.0 [0.8,1.1] | 1.2 [0.9,1.6] | 3.1E-01 |
| early-only SSRI exposure (582) | late SSRI exposure (2133) | 1.5 [1.0,2.3]  + | 1.3 [0.8,2.1] | 2.2E-01 | 1.2 [0.9,1.6] | 1.2 [0.9,1.6] | 3.3E-01 |
| no SSRI exposure (2350) | any SSRI exposure (2715) | 1.2 [1.0,1.6] | 1.9 [1.3,2.8] ** | 1.2E-03 | 0.9 [0.8,1.1] | 1.1 [0.9,1.5] | 3.5E-01 |

| exposure status to psychotropic medication as an additional covariate^e^ | | | | | | | |
| --- | --- | --- | --- | --- | --- | --- | --- |
| reference group (n) | comparison group (n) | unadjusted PTB OR | adjusted PTB OR | adjusted PTB OR p-value | unadjusted GA β | adjusted GA β | adjusted GA β p-value |
| no SSRI exposure (3122) | early-only SSRI exposure (690) | 0.8 [0.5,1.0]  + | 1.4 [0.8,2.4] | 2.3E-01 | 1.1 [0.0,2.1] * | -0.3 [-1.9,1.4] | 7.6E-01 |
| no-late SSRI exposure (3812) | late SSRI exposure (2596) | 1.4 [1.2,1.7] **** | 1.6 [1.3,1.9] **** | 3.6E-05 | -1.9 [-2.6,-1.3] ***** | -2.3 [-3.1,-1.5] ***** | 1.2E-08 |
| no SSRI exposure (3122) | late SSRI exposure (2596) | 1.4 [1.1,1.6] *** | 1.7 [1.3,2.1] *** | 1.0E-04 | -1.8 [-2.4,-1.1] ***** | -2.6 [-3.6,-1.6] ***** | 4.5E-07 |
| early-only SSRI exposure (690) | late SSRI exposure (2596) | 1.8 [1.3,2.5] *** | 1.5 [1.1,2.1] * | 1.2E-02 | -2.8 [-3.9,-1.7] ***** | -1.9 [-3.0,-0.8] *** | 8.5E-04 |
| no SSRI exposure (3122) | any SSRI exposure (3286) | 1.2 [1.0,1.5] * | 1.6 [1.2,2.1] *** | 3.4E-04 | -1.2 [-1.8,-0.5] *** | -2.3 [-3.3,-1.3] ***** | 3.9E-06 |

| exposure status to psychotropic medication as an additional covariate^e^ (continued) | | | | | | | |
| --- | --- | --- | --- | --- | --- | --- | --- |
| reference group (n) | comparison group (n) | unadjusted LBW OR | adjusted LBW OR | adjusted PTB OR p-value | unadjusted SGA OR | adjusted SGA OR | adjusted SGA OR p-value |
| no SSRI exposure (3122) | early-only SSRI exposure (690) | 0.7 [0.4,1.0]+ | 1.0 [0.5,2.0] | 9.6E-01 | 0.8 [0.6,1.0]+ | 1.0 [0.6,1.6] | 9.8E-01 |
| no-late SSRI exposure (3812) | late SSRI exposure (2596) | 1.3 [1.1,1.6] ** | 1.7 [1.3,2.2] *** | 1.0E-04 | 1.0 [0.9,1.2] | 1.2 [1.0,1.5]+ | 6.6E-02 |
| no SSRI exposure (3122) | late SSRI exposure (2596) | 1.3 [1.0,1.6] * | 1.7 [1.3,2.4] *** | 5.5E-04 | 1.0 [0.8,1.1] | 1.2 [0.9,1.5] | 2.0E-01 |
| early-only SSRI exposure (690) | late SSRI exposure (2596) | 1.9 [1.2,2.9] ** | 1.6 [1.1,2.5] * | 2.4E-02 | 1.3 [1.0,1.7] | 1.2 [0.9,1.6] | 2.0E-01 |
| no SSRI exposure (3122) | any SSRI exposure (3286) | 1.1 [0.9,1.4] | 1.7 [1.2,2.3] ** | 1.5E-03 | 0.9 [0.8,1.1] | 1.2 [0.9,1.5] | 2.0E-01 |

| exclusion of women with single SSRI prescription during pregnancy^f^ | | | | | | | |
| --- | --- | --- | --- | --- | --- | --- | --- |
| reference group (n) | comparison group (n) | unadjusted PTB OR | adjusted PTB OR | adjusted PTB OR p-value | unadjusted GA β | adjusted GA β | adjusted GA β p-value |
| no SSRI exposure (3122) | early-only SSRI exposure (96) | 1.0 [0.5,2.0] | 1.2 [0.5,3.0] | 7.4E-01 | 0.9 [-1.7,3.4] | -0.5 [-3.8,2.7] | 7.4E-01 |
| no-late SSRI exposure (3218) | late SSRI exposure (1378) | 1.6 [1.3,1.9] ***** | 1.8 [1.4,2.4] **** | 3.3E-05 | -2.6 [-3.4,-1.8] ***** | -3.1 [-4.3,-2.0] ***** | 9.0E-08 |
| no SSRI exposure (3122) | late SSRI exposure (1378) | 1.6 [1.3,1.9] **** | 1.9 [1.4,2.5] **** | 1.9E-05 | -2.6 [-3.4,-1.7] ***** | -3.2 [-4.4,-2.0] ***** | 2.6E-07 |
| early-only SSRI exposure (96) | late SSRI exposure (1378) | 1.6 [0.8,3.4] | 1.4 [0.7,3.1] | 3.7E-01 | -3.4 [-6.3,-0.5] * | -2.3 [-5.4,0.7] | 1.4E-01 |
| no SSRI exposure (3122) | any SSRI exposure (1474) | 1.5 [1.3,1.9] **** | 1.9 [1.4,2.5] **** | 3.0E-05 | -2.3 [-3.2,-1.5] ***** | -3.1 [-4.3,-1.9] ***** | 3.6E-07 |

| exclusion of women with single SSRI prescription during pregnancy^f^ (continued) | | | | | | | |
| --- | --- | --- | --- | --- | --- | --- | --- |
| reference group (n) | comparison group (n) | unadjusted LBW OR | adjusted LBW OR | adjusted PTB OR p-value | unadjusted SGA OR | adjusted SGA OR | adjusted SGA OR p-value |
| no SSRI exposure (3122) | early-only SSRI exposure (96) | 0.6 [0.2,1.8] | 0.7 [0.2,2.6] | 5.4E-01 | 0.8 [0.4,1.6] | 0.9 [0.4,2.1] | 8.4E-01 |
| no-late SSRI exposure (3218) | late SSRI exposure (1378) | 1.4 [1.1,1.8] ** | 1.9 [1.3,2.7] *** | 3.4E-04 | 1.0 [0.8,1.2] | 1.2 [0.9,1.5] | 2.0E-01 |
| no SSRI exposure (3122) | late SSRI exposure (1378) | 1.4 [1.1,1.8] * | 1.9 [1.3,2.7] *** | 6.2E-04 | 1.0 [0.8,1.2] | 1.2 [0.9,1.6] | 2.2E-01 |
| early-only SSRI exposure (96) | late SSRI exposure (1378) | 2.5 [0.8,8.0] | 2.3 [0.7,7.5] | 1.7E-01 | 1.2 [0.6,2.3] | 1.2 [0.6,2.5] | 5.7E-01 |
| no SSRI exposure (3122) | any SSRI exposure (1474) | 1.3 [1.0,1.7] * | 1.8 [1.3,2.6] ** | 1.1E-03 | 1.0 [0.8,1.2] | 1.2 [0.9,1.6] | 2.1E-01 |

**Table S7 Result of other antidepressants analyses (N06AA, N06AX)**

| N06AA exposure | | | | | | | |
| --- | --- | --- | --- | --- | --- | --- | --- |
| reference group (n) | comparison group (n) | unadjusted PTB OR | adjusted PTB OR | adjusted PTB OR p-value | unadjusted GA β | adjusted GA β | adjusted GA β p-value |
| no N06AA exposure (3015) | early-only N06AA exposure (91) | 0.9 [0.4,1.9] | 0.6 [0.2,2.1] | 4.2E-01 | -1.4 [-4.1,1.2] | -0.6 [-5.2,4.0] | 7.9E-01 |
| no-late N06AA exposure (3106) | late N06AA exposure (83) | 2.2 [1.2,3.9] * | 2.0 [0.9,4.2]+ | 7.5E-02 | -5.5 [-8.3,-2.7] *** | -3.6 [-6.9,-0.2] * | 3.8E-02 |
| no N06AA exposure (3015) | late N06AA exposure (83) | 2.1 [1.2,3.9] * | 1.6 [0.6,4.0] | 3.5E-01 | -5.5 [-8.2,-2.7] **** | -3.1 [-7.1,0.9] | 1.3E-01 |
| early-only N06AA exposure (91) | late N06AA exposure (83) | 2.4 [0.9,6.4]+ | 4.5 [1.4,14.5] * | 1.2E-02 | -4.1 [-9.2,1.1] | -7.0 [-13.2,-0.9] * | 2.6E-02 |
| no N06AA exposure (3015) | any N06AA exposure (174) | 1.5 [0.9,2.3] | 1.0 [0.4,2.3] | 9.6E-01 | -3.4 [-5.3,-1.4]  *** | -1.6 [-5.0,1.8] | 3.5E-01 |

| N06AA exposure (continued) | | | | | | | |
| --- | --- | --- | --- | --- | --- | --- | --- |
| reference group (n) | comparison group (n) | unadjusted LBW OR | adjusted LBW OR | adjusted LBW OR p-value | unadjusted SGA OR | adjusted SGA OR | adjusted SGA OR p-value |
| no N06AA exposure (3015) | early-only N06AA exposure (91) | 1.2 [0.5,2.8] | 1.5 [0.3,6.8] | 6.1E-01 | 1.4 [0.8,2.5] | 1.2 [0.4,3.2] | 7.8E-01 |
| no-late N06AA exposure (3106) | late N06AA exposure (83) | 2.5 [1.3,4.9] ** | 2.6 [1.1,6.0] * | 3.0E-02 | 1.2 [0.6,2.2] | 1.1 [0.5,2.4] | 7.3E-01 |
| no N06AA exposure (3015) | late N06AA exposure (83) | 2.5 [1.3,4.9] ** | 2.5 [0.9,7.1]+ | 8.2E-02 | 1.2 [0.6,2.2] | 1.7 [0.7,4.1] | 2.5E-01 |
| early-only N06AA exposure (91) | late N06AA exposure (83) | 2.1 [0.7,6.0] | 3.1 [0.9,10.8]+ | 8.1E-02 | 0.8 [0.4,1.9] | 0.6 [0.2,1.5] | 2.4E-01 |
| no N06AA exposure (3015) | any N06AA exposure (174) | 1.8 [1.1,3.1] * | 2.1 [0.8,5.4] | 1.3E-01 | 1.3 [0.9,2.0] | 1.5 [0.7,3.1] | 3.1E-01 |

| N06AX exposure | | | | | | | |
| --- | --- | --- | --- | --- | --- | --- | --- |
| reference group (n) | comparison group (n) | unadjusted PTB OR | adjusted PTB OR | adjusted PTB OR p-value | unadjusted GA β | adjusted GA β | adjusted GA β p-value |
| no N06AX exposure (2434) | early-only N06AX exposure (377) | 1.3 [0.9,1.9] | 1.1 [0.6,2.2] | 7.8E-01 | -0.6 [-2.0,0.7] | 0.5 [-1.9,2.9] | 6.7E-01 |
| no-late N06AX exposure (2811) | late N06AX exposure (830) | 1.7 [1.3,2.2] **** | 1.7 [1.2,2.3] ** | 1.1E-03 | -2.6 [-3.6,-1.7] ***** | -2.5 [-3.8,-1.3] **** | 6.6E-05 |
| no N06AX exposure (2434) | late N06AX exposure (830) | 1.7 [1.4,2.3] **** | 2.3 [1.5,3.6] **** | 5.9E-05 | -2.7 [-3.7,-1.7] ***** | -3.3 [-5.0,-1.6] *** | 1.1E-04 |
| early-only N06AX exposure (377) | late N06AX exposure (830) | 1.3 [0.9,2.0] | 1.4 [0.9,2.1] | 1.5E-01 | -2.1 [-3.8,-0.4] * | -2.3 [-4.1,-0.6] ** | 9.6E-03 |
| no N06AX exposure (2434) | any N06AX exposure (1207) | 1.6 [1.3,2.0] **** | 2.1 [1.4,3.1] *** | 3.9E-04 | -2.1 [-3.0,-1.2] ***** | -2.4 [-4.0,-0.9] ** | 2.5E-03 |

| N06AX exposure (continued) | | | | | | | |
| --- | --- | --- | --- | --- | --- | --- | --- |
| reference group (n) | comparison group (n) | unadjusted LBW OR | adjusted LBW OR | adjusted LBW OR p-value | unadjusted SGA OR | adjusted SGA OR | adjusted SGA OR p-value |
| no N06AX exposure (2434) | early-only N06AX exposure (377) | 1.1 [0.7,1.7] | 1.1 [0.4,2.5] | 9.0E-01 | 0.8 [0.6,1.2] | 0.8 [0.4,1.5] | 5.0E-01 |
| no-late N06AX exposure (2811) | late N06AX exposure (830) | 1.7 [1.2,2.3] *** | 2.1 [1.4,3.1] *** | 1.7E-04 | 1.3 [1.0,1.6] * | 1.5 [1.1,2.0] ** | 9.4E-03 |
| no N06AX exposure (2434) | late N06AX exposure (830) | 1.7 [1.2,2.3] *** | 2.8 [1.7,4.5] **** | 4.1E-05 | 1.3 [1.0,1.6] * | 1.4 [1.0,2.1]+ | 6.3E-02 |
| early-only N06AX exposure (377) | late N06AX exposure (830) | 1.6 [0.9,2.7]+ | 1.5 [0.9,2.6] | 1.3E-01 | 1.5 [1.0,2.3] * | 1.6 [1.0,2.3] * | 3.3E-02 |
| no N06AX exposure (2434) | any N06AX exposure (1207) | 1.5 [1.1,2.0] ** | 2.5 [1.5,4.0] *** | 1.9E-04 | 1.1 [0.9,1.4] | 1.3 [0.9,1.9] | 1.7E-01 |

| SNRI exposure | | | | | | | |
| --- | --- | --- | --- | --- | --- | --- | --- |
| reference group (n) | comparison group (n) | unadjusted PTB OR | adjusted PTB OR | adjusted PTB OR p-value | unadjusted GA β | adjusted GA β | adjusted GA β p-value |
| no SNRI exposure (2434) | early-only SNRI exposure (371) | 1.3 [0.9,1.9] | 1.1 [0.6,2.2] | 7.8E-01 | -0.6 [-2.0,0.7] | 0.5 [-1.9,2.9] | 6.7E-01 |
| no-late SNRI exposure (2811) | late SNRI exposure (823) | 1.7 [1.3,2.2] **** | 1.8 [1.3,2.4] *** | 6.1E-04 | -2.6 [-3.6,-1.7] ***** | -2.5 [-3.8,-1.3] **** | 6.6E-05 |
| no SNRI exposure (2434) | late SNRI exposure (823) | 1.8 [1.4,2.3] **** | 2.4 [1.6,3.6] **** | 5.9E-05 | -2.7 [-3.7,-1.7] ***** | -3.3 [-5.0,-1.6] *** | 1.1E-04 |
| Early-only SNRI exposure (377) | late SNRI exposure (823) | 1.4 [0.9,2.1] | 1.4 [0.9,2.1] | 1.1E-01 | -2.1 [-3.8,-0.4] * | -2.3 [-4.1,-0.6] ** | 9.6E-03 |
| no SNRI exposure (2434) | any SNRI exposure (1194) | 1.6 [1.3,2.0] **** | 2.1 [1.4,3.1] *** | 3.2E-04 | -2.2 [-3.0,-1.2] ***** | -2.5 [-4.0,-1.0] ** | 1.7E-03 |

| SNRI exposure (continued) | | | | | | | |
| --- | --- | --- | --- | --- | --- | --- | --- |
| reference group (n) | comparison group (n) | unadjusted LBW OR | adjusted LBW OR | adjusted LBW OR p-value | unadjusted SGA OR | adjusted SGA OR | adjusted SGA OR p-value |
| no SNRI exposure (2434) | early-only SNRI exposure (371) | 1.1 [0.7,1.7] | 1.1 [0.4,2.5] | 9.0E-01 | 0.8 [0.6,1.2] | 0.8 [0.4,1.5] | 5.0E-01 |
| no-late SNRI exposure (2811) | late SNRI exposure (823) | 1.7 [1.2,2.3] *** | 2.1 [1.4,3.1] *** | 1.6E-04 | 1.3 [1.0,1.6] * | 1.5 [1.1,2.0] ** | 8.0E-03 |
| no SNRI exposure (2434) | late SNRI exposure (823) | 1.7 [1.2,2.3] *** | 2.8 [1.7,4.5] **** | 4.0E-05 | 1.3 [1.0,1.6] * | 1.5 [1.0,2.1]* | 6.3E-02 |
| early-only SNRI exposure (377) | late SNRI exposure (823) | 1.6 [0.9,2.7]+ | 1.5 [0.9,2.6] | 1.2E-01 | 1.5 [1.0,2.3] * | 1.6 [1.0,2.4] * | 3.3E-02 |
| no SNRI exposure (2434) | any SNRI exposure (1194) | 1.5 [1.1,2.0] ** | 2.5 [1.5,4.0] *** | 1.9E-04 | 1.1 [0.9,1.4] | 1.3 [0.9,1.9] | 1.4E-01 |

**Table S8 Variable definition**

| Variable Definition | | |
| --- | --- | --- |
| Category | Feature | Definition |
| pregnancy characteristics | parity | number of times a women has given birth to a fetus older than 24 weeks of gestational age; 0, 1~4, ≥5 |
|  | preterm history | number of times a women has given preterm birth |
|  | delivery year | year of delivery; 2013, 2014, 2015, 2016, 2017, 2018, 2019, 2020 |
|  | fetal sex | fetal sex; female, male |
| maternal characteristics | age at LMP | maternal age at the start of pregnancy |
|  | racial group | maternal race group; white/caucasian, black/african american, asian, american indian, alaska native, native hawaiian/other pacific islanders, multirace, unknown/not reported |
|  | ethnic group | maternal ethnicity group; hispanic/latino, not hispanic/latino, unknown/not reported |
|  | insurance status | insurance status; commercial, medicaid/medicare |
|  | pregravid BMI category | pregravid body mass index (BMI); underweight, normal, overweight, obese |
|  | smoking status | smoking history reported during prenatal visit; 1,0 |
|  | illegal drug use status | illegal drug use history reported during prenatal visit; 1,0 |
|  | alcohol drinking status | alcohol drinking status reported during prenatal visit; 1,0 |
| social vulnerability | social vulnerability index | social vulnerability percentile ranking of women’s census tract; 0~1 |
| comorbidities | anemia | any diagnosis of anemia or descendent disorders during two-year pre-pregnancy period; 1,0 |
|  | asthma | any diagnosis of asthma or descendent disorders during two-year pre-pregnancy period; 1,0 |
|  | chronic lung disease | any diagnosis of chronic lung disease or descendent disorders during two-year pre-pregnancy period; 1,0 |
|  | cardiovascular disease | any diagnosis of cardiovascular disease or descendent disorders during two-year pre-pregnancy period; 1,0 |
|  | cystic fibrosis | any diagnosis of cystic fibrosis or descendent disorders during two-year pre-pregnancy period; 1,0 |
|  | diabetes | any diagnosis of diabetes or descendent disorders during two-year pre-pregnancy period; 1,0 |
|  | leukemia | any diagnosis of leukemia or descendent disorders during two-year pre-pregnancy period; 1,0 |
|  | pneumonia | any diagnosis of pneumonia or descendent disorders during two-year pre-pregnancy period; 1,0 |
|  | renal diseases | any diagnosis of renal diseases or descendent disorders during two-year pre-pregnancy period; 1,0 |
|  | sepsis | any diagnosis of sepsis or descendent disorders during two-year pre-pregnancy period; 1,0 |
|  | sickle cell diseases | any diagnosis of sickle cell diseases or descendent disorders during two-year pre-pregnancy period; 1,0 |

| Variable Definition (continued) | | |
| --- | --- | --- |
| Category | Feature | Definition |
| pre-pregnancy mental condition | adjustment disorder | any diagnosis of adjustment disorder or descendent disorders during 6-month pre-pregnancy period; 1,0 |
|  | anxiety disorder | any diagnosis of anxiety disorder or descendent disorders during 6-month pre-pregnancy period; 1,0 |
|  | PHQ-9 score | latest patient health questionnaire-9 (PHQ-9) score during two-year pre-pregnancy period |
|  | PHQ-9 category | category of PHQ-9 score; minimal: 1-4, mild: 5-9, moderate: 10-14, moderately severe: 15-19, severe: 20-27 |
|  | N06AA exposure | any prescription order of N06AA during 6-month pre-pregnancy period; 1,0 |
|  | N06AB exposure | any prescription order of SSRI(N06AB) during 6-month pre-pregnancy period; 1,0 |
|  | N06AG exposure | any prescription order of N06AG during 6-month pre-pregnancy period; 1,0 |
|  | N06AF exposure | any prescription order of N06AF during 6-month pre-pregnancy period; 1,0 |
|  | N06AX exposure | any prescription order of N06AX during 6-month pre-pregnancy period; 1,0 |
| Prenatal  mental condition | late pregnancy depression diagnosis | any depression diagnosis during second or third trimester; 1,0 |
|  | PHQ-4 score | latest patient health questionnaire-4 (PHQ-4) score during two-year pregnancy |
|  | PHQ-4 category | category of PHQ-4 score; normal: 0-2, mild:3-5, moderate: 6-8, severe: 9-12 |

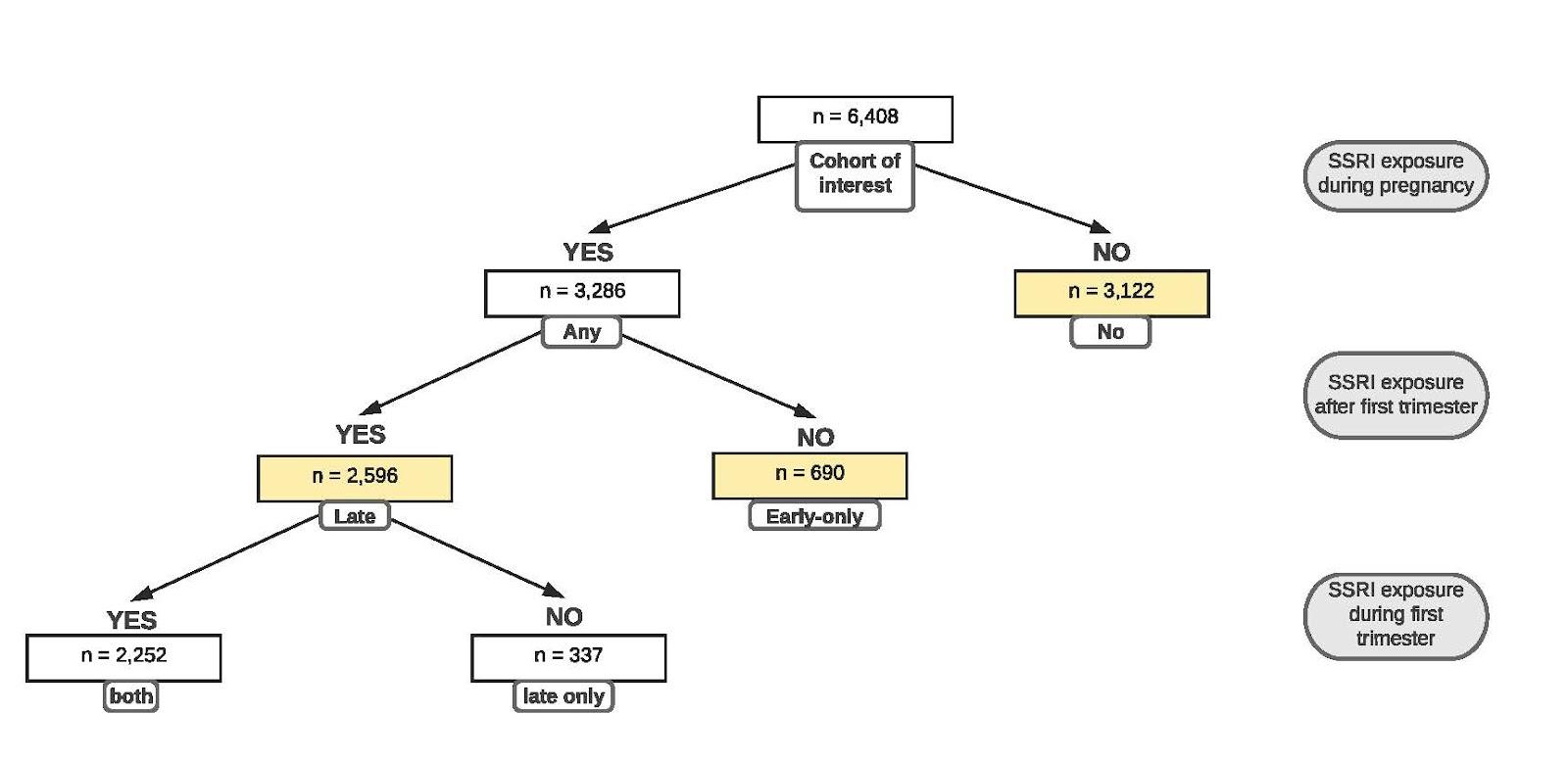

**Figure S1:** Flow diagram of exposure group selection

Abbreviation: SSRI, Selective Serotonin Reuptake Inhibitor.
